# Synovial Monocytes Drive the Pathogenesis in Oligoarticular Juvenile Idiopathic Arthritis via IL-6/JAK/STAT Signalling and Cell-Cell Interactions

**DOI:** 10.1101/2023.01.17.23284466

**Authors:** Tobias Schmidt, Alma Dahlberg, Elisabet Berthold, Petra Król, Sabine Arve-Butler, Emilia Rydén, Seyed Morteza Najibi, Anki Mossberg, Anders Bengtsson, Fredrik Kahn, Bengt Månsson, Robin Kahn

**Affiliations:** Department of Pediatrics, Lund University, Sweden; Department of Rheumatology, Lund University, Sweden; Department of Infection medicine, Clinical Sciences Lund, Lund University, Sweden; Wallenberg Center for Molecular Medicine, Lund University, Sweden

**Keywords:** Monocytes, juvenile idiopathic arthritis, inflammation, synovial fluid, IL-6, STAT3

## Abstract

**Objectives:** Synovial monocytes in oligoarticular juvenile idiopathic arthritis (oJIA) are polarized, but little is known of how they contribute to disease and attain their pathogenic features. The aim of this study was to investigate the role of monocytes in the pathogenesis of oJIA.

**Methods:** The function of synovial monocytes was analysed by several assays believed to reflect key pathogenic events, such as T-cell activation-, efferocytosis- and cytokine production assays through flow cytometry in untreated oJIA patients (n=33). The effect of synovial fluid on healthy monocytes was investigated through mass spectrometry, broad-spectrum phosphorylation assays and functional assays. Additional effects on monocytes were studied through co-cultures with primary fibroblast-like synoviocytes.

**Results:** The results demonstrate that synovial monocytes display functional alterations, e.g., increased ability to induce T-cell activation, increased efferocytosis and resistance to cytokine production following activation with LPS. *In vitro*, synovial fluid induced regulatory features in healthy monocytes through an IL-6/JAK/STAT mechanism. The magnitude of synovial IL-6 driven activation in monocytes was reflected in circulating cytokine levels. An increased ability to induce T-cell activation and markers of antigen presentation could be induced by co-culture with fibroblast-like synoviocytes.

**Conclusions:** Synovial monocytes in oJIA are functionally affected, drive chronic inflammation, and promote adaptive immune responses. This phenotype can be replicated *in vitro* through a combination of synovial fluid (through IL-6/JAK/STAT) and cell-cell interactions. These data support a role of monocytes in the pathogenesis of oJIA and highlight a group of patients more likely to benefit from targeting the IL-6/JAK/STAT axis to restore synovial homeostasis.

**Key messages:** *What is already known on this topic:*

- Monocytes infiltrate the joint in oligoarticular juvenile idiopathic arthritis (JIA), where they display a pathogenic phenotype and signs of activation

*What this study adds:*

- The results of this study demonstrate functional alterations of synovial monocytes in driving chronic inflammation in oligoarticular JIA
- Synovial monocytes acquire their regulatory properties through the IL-6/JAK/STAT pathway in synovial fluid and their inflammatory properties through cell-cell interactions
- In patients with high IL-6/JAK/STAT involvement, this is reflected in elevated circulating cytokine levels

*How this study might affect research, practice or policy:*

- This study describes the mechanisms controlling the function of synovial monocytes in oligoarticular JIA and identifies patients likely to respond to IL-6/JAK/STAT inhibition, which should be further explored to facilitate personalized medicine.

## Introduction

Juvenile idiopathic arthritis (JIA) is the most common rheumatic disease in children, mainly affecting the joints. It is defined as persistent arthritis of unknown origin with an onset before the age of 16 (1). JIA is a heterogenous disease that includes seven different subtypes. The most common one in the western world is oligoarticular JIA (oJIA), which accounts for 30-60% of all patients (2, 3). These patients often display asymmetric arthritis, in one to four joints, and it is believed to be a distinct phenotype rarely seen in adult arthritis. The children usually present at preschool age, are antinuclear-antibody (ANA) positive and have a high risk of developing uveitis (2). However, the pathogenesis of oJIA is largely unknown. The affected joint is characterized by hyperplasia, inflammation, and presence of infiltrating immune cells from both the innate and adaptive immune system (4). T- and B-cells are thought to play a role, given the genetic relationship to MHC class II alleles and the presence of ANAs (4-6). Though, the contribution of members from the innate immune system, such as monocytes, is becoming increasingly recognized (7-10).

Monocytes are key players in the innate immune system, with roles in antigen presentation, cytokine production and phagocytosis. They are found in the synovial fluid of patients with adult arthritis, displaying several markers of activation compared to circulating monocytes, such as CD16, HLA-DR and toll-like receptors (TLRs) (11, 12). Furthermore, synovial monocytes and macrophages display a state of polarization (13). Polarization, or activation, is the process in which these cells respond to the environment and acquire distinct phenotypes, a process that also influences the cells’ effector functions (14). Traditionally, these are classified as pro-inflammatory or anti-inflammatory/regulatory phenotypes, although polarization most likely represents a continuum were these cells display features from both endpoints. Multiple cytokines found within the inflamed joint can induce polarization, which mainly signals through the JAK/STAT pathway (13, 14). Interestingly, polarization may differ between different forms of arthritis, which may have implications in the function of these cells in different arthritides (15, 16). Synovial monocytes and macrophages are believed to drive pro-inflammatory processes via cell-cell interactions, e.g., with fibroblast-like synoviocytes and T-cells, and production of soluble mediators such as cytokines (17, 18). In addition, features of regulatory monocytes and macrophages, such as angiogenesis, can facilitate chronic inflammation in arthritis (19, 20). CD163, a classic marker of regulatory monocytes and macrophages, correlate with inflammation in spondylarthritis (21). Finally, these cells can also be potent sources of inflammatory cytokines (22). Thus, both inflammatory and regulatory polarization may be detrimental in chronic inflammation.

The role of monocytes in oJIA is poorly characterized compared to adult arthritis. We and others have previously shown that synovial monocytes from patients with oJIA are polarized, with both inflammatory and regulatory features (7, 23). Monocytes and macrophages are believed to contribute to the pathogenesis in a similar fashion as adult arthritides, such as through cytokine production (24), but extensive data on the role of monocytes in oJIA is lacking. Here, we set out to investigate how synovial monocytes are functionally affected and contribute to the pathogenesis of oJIA, the mechanisms and pathways driving this phenotype, and the potential impact of these pathways in the clinical setting.

## Materials and Methods

Details and descriptions on sample preparation, methodology and assays can be found in **supplementary Materials and Methods**.

### Patient material and clinical characteristics

Patients (n=33) with oligoarticular JIA (oJIA) according to the International League of Associations for Rheumatology (ILAR) at the Department of Paediatric Rheumatology, Skåne University Hospital, Sweden between 2016-2022, were included in this study when undergoing therapeutic joint aspiration. This study was approved by the Regional Ethical Review Board for southern Sweden (2016/128). Patients were included upon written informed consent from the patients and/or their legal guardians. The patients had not received any disease-modifying anti-rheumatic drugs (DMARDs) or steroids, neither oral nor intra-articular, in the last 6 months prior to inclusion, but may have been administered non-steroid anti-inflammatory drugs (NSAIDs). The patient characteristics are described in **Table 1**. Synovial fluid (SF) and blood (in either EDTA-, serum-, or heparin tubes (BD Biosciences)) were collected. Cytokines (IL-6, IL-8, CRP, SAA, TNF and IFNα2a) were analysed in SF or plasma samples (Mesoscale Diagnostics).

**Table 1.**
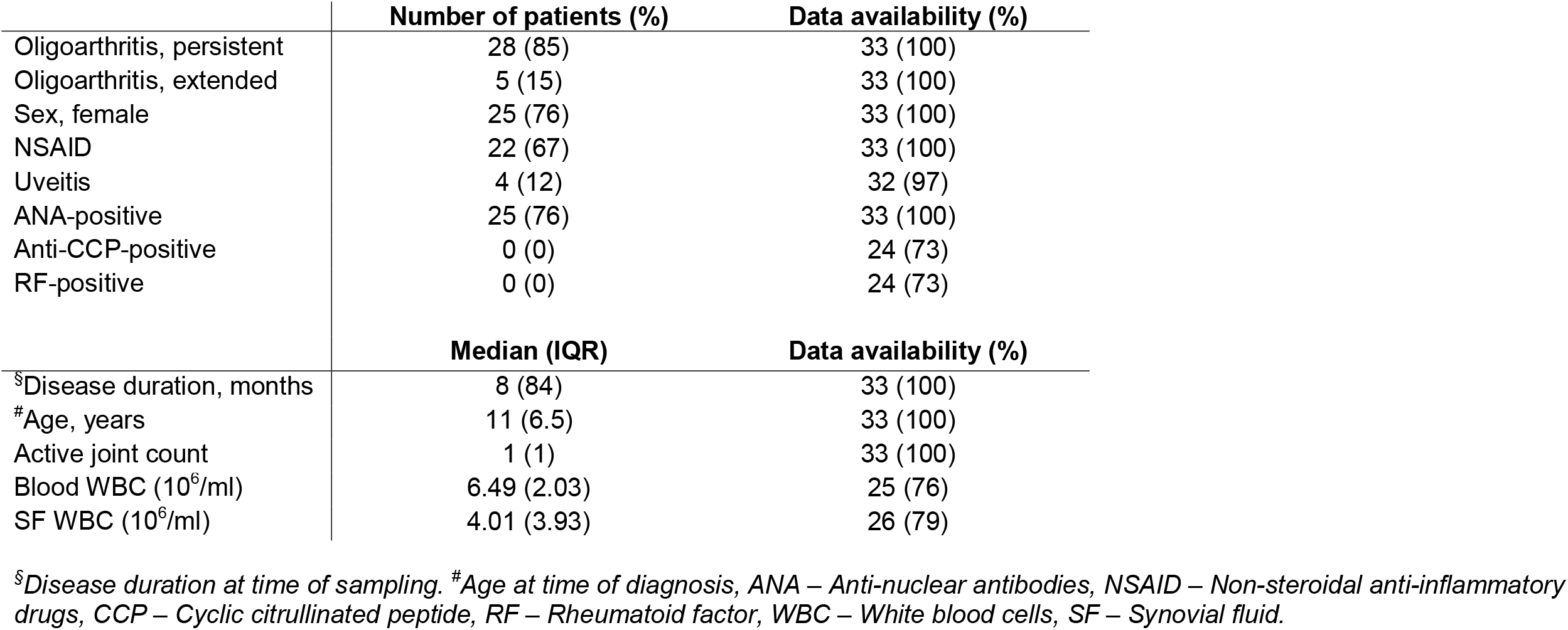
Overview of the patient cohort. Clinical and laboratory data of the 33 patients included in this study.

|  | Number of patients (%) | Data availability (%) |
| --- | --- | --- |
| Oligoarthritis, persistent | 28 (85) | 33 (100) |
| Oligoarthritis, extended | 5 (15) | 33 (100) |
| Sex, female | 25 (76) | 33 (100) |
| NSAID | 22 (67) | 33 (100) |
| Uveitis | 4 (12) | 32 (97) |
| ANA-positive | 25 (76) | 33 (100) |
| Anti-CCP-positive | 0 (0) | 24 (73) |
| RF-positive | 0 (0) | 24 (73) |
|  | Median (IQR) | Data availability (%) |
| <sup>§</sup>Disease duration, months | 8 (84) | 33 (100) |
| <sup>#</sup>Age, years | 11 (6.5) | 33 (100) |
| Active joint count | 1 (1) | 33 (100) |
| Blood WBC (10<sup>6</sup>/ml) | 6.49 (2.03) | 25 (76) |
| SF WBC (10<sup>6</sup>/ml) | 4.01 (3.93) | 26 (79) |

<sup>§</sup>Disease duration at time of sampling. <sup>#</sup>Age at time of diagnosis, ANA – Anti-nuclear antibodies, NSAID – Non-steroidal anti-inflammatory drugs, CCP – Cyclic citrullinated peptide, RF – Rheumatoid factor, WBC – White blood cells, SF – Synovial fluid.

### Patient and public involvement

Patient partners from the Swedish League Against Rheumatism have been involved in the design and dissemination of the study. During the initiation, we discussed with patient partners on the relevance of the project as well as the information to participants and frequency/amount of sampling. Once the project was ongoing, our study has been supported financially and featured in the yearly report from research foundation of the Swedish League Against Rheumatism.

### Monocyte isolation, polarization and culture

Monocytes were isolated from healthy controls upon informed consent using magnetic CD14^+^ bead isolation according to the manufacturer’s instructions (Miltenyi). Monocytes from healthy controls at 1×10^6^/ml in RPMI-1640 medium were polarized overnight with 20% SF (or paired serum as control) to generate *in vitro* polarized monocytes. The influence of fibroblast-like synoviocytes (FLS) and migration on the monocyte phenotype was analysed by co-culture- and transwell systems, respectively.

### Flow cytometry analysis, phosphorylation assay and proteomics

Monocytes from patients and *in vitro* polarized monocytes were analysed for surface marker expression, efferocytosis, T-cell activation, STAT phosphorylation, and cytokine production by flow cytometry. *In vitro* polarized monocytes were also analysed for phagocytosis and ROS production. Tofacitinib and tocilizumab were used to block IL-6/JAK/STAT signalling and their effect on viability was considered minor (**supplementary figure 1**). Gating strategies can be found in **supplementary figure 2**. In addition, *in vitro* polarized monocytes were analysed by broad-spectrum phosphorylation assay and liquid chromatography mass spectrometry.

## Results

### Synovial monocytes from oJIA patients display pathogenic alterations, such as stimulation of T-cell activation, increased efferocytosis, and show signs of previous activation

To investigate if monocytes from the joints of oJIA patients are functionally affected, we obtained synovial fluid (SF) and blood, to compare synovial monocytes to paired circulating monocytes (**figure 1A**). When co-cultured with healthy T-cells, synovial monocytes displayed an increased ability to induce proliferation (p=0.0068) and activation markers in T-cells (CD25 (p=0.0312), HLA and CTLA-4 (both p=0.0156)) compared to circulating monocytes (**figure 1B and C**). In parallel, synovial monocytes had increased expression of markers related to antigen presentation (CD86 (p<0.0001) and HLA, (p=0.0078, **figure 1D**). Thus, synovial monocytes may contribute to prolongating synovial inflammation through linking innate immunity with adaptive immunity.

**Figure 1.**
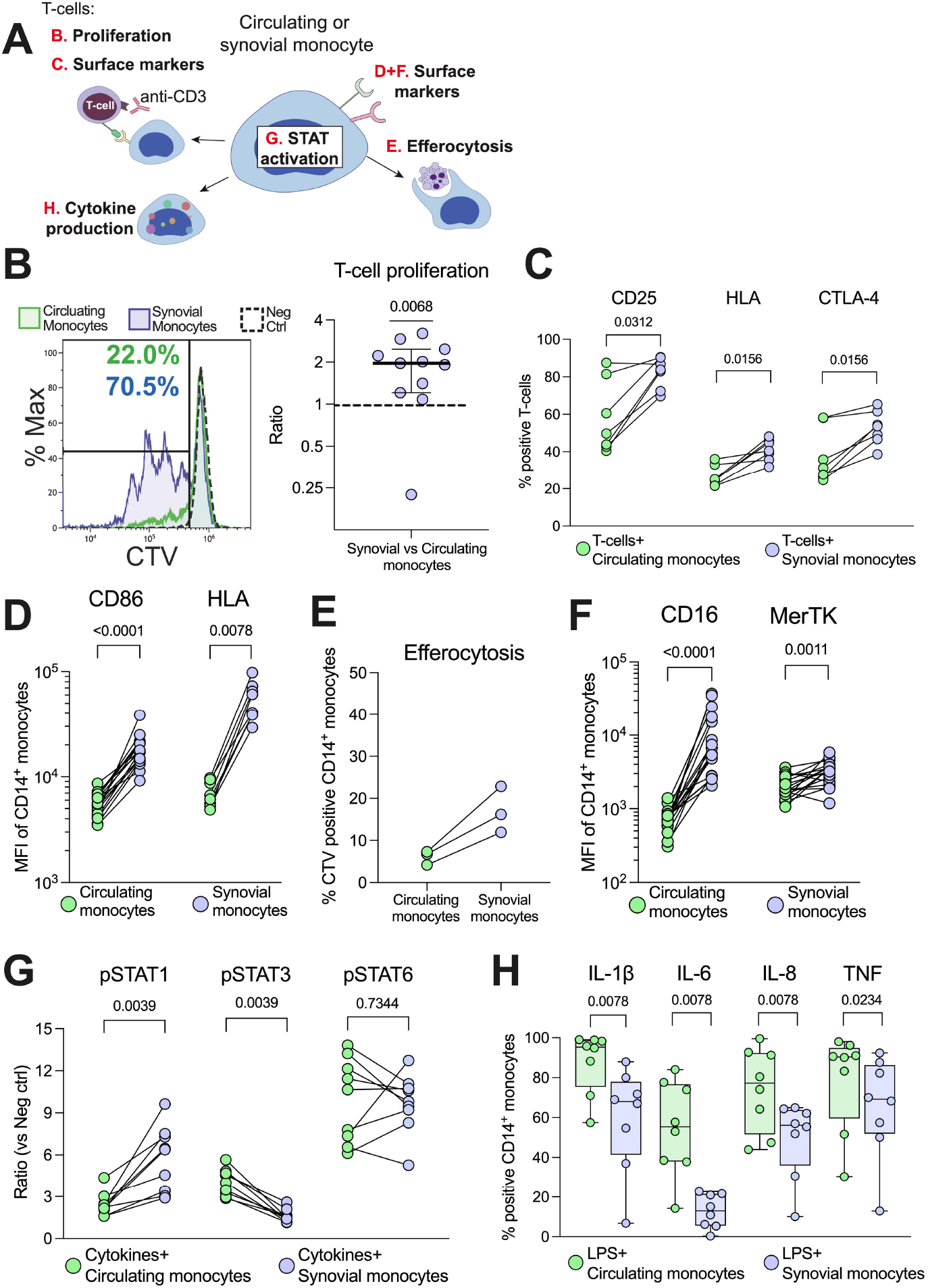
Synovial monocytes from patients with oJIA induce T-cell proliferation, display increased efferocytosis and a resistance to further activation. (**A**) Schematic overview of the various analyses of synovial- vs circulating monocytes. (**B**) Isolated monocytes were incubated with CTV-stained CD3-activated healthy T-cells (1:10 monocytes to T-cells) for 72hrs, which were subsequently analyzed by flow cytometry for proliferation (presented as the ratio of percent proliferation induced by synovial-vs circulating monocytes, line at median, n=11) and (**C**) expression of activation markers on T-cells (n=7). (**D**) Shows expression of markers related to antigen presentation on monocytes: CD86 (n=17) and HLA (n=8). (**E**) As a measurement of clearance, isolated monocytes were incubated with CTV-stained apoptotic neutrophils for 3hrs (n=3), and the percent of CTV-positive monocytes is presented (reflecting uptake of neutrophils). Monocytes were defined as CD14^+^CD66b^-^. (**F**) Shows expression of markers related to clearance on monocytes (CD16 and MerTK, n=17). (**G**) Monocytes in synovial fluid or blood were activated with IFNγ, IL-4 and IL-6 for 15min, and investigated for their phosphorylation response of STAT1, 3 or 6 (n=9). Values represent the ratio between stimulated vs unstimulated samples. (**H**) Cytokine production of monocytes was studied intracellularly by incubation with golgiplug, followed by LPS activation for 4hrs (n=8). Statistics were performed using one sample Wilcoxon signed-rank test (with the hypothetical median of 1) or Wilcoxon matched pairs signed rank test. *oJIA-Oligoarticular juvenile idiopathic arthritis, MFI-Median fluorescence intensity, IFN-Interferon, IL-Interleukin, STAT-Signal transducer and activator of transcription, CTV-Cell Trace Violet, LPS-Lipopolysaccharide, TNF-Tumor necrosis factor*.

Patient synovial monocytes also displayed increased efferocytosis of apoptotic neutrophils (n=3, **figure 1E**), as well as increased expression of surface markers related to clearance (CD16, p<0.0001 and MerTK, p=0.0011 **figure 1F**). Hence, synovial monocytes also display regulatory mechanisms of increased clearance of apoptotic material. Finally, we investigated how patient synovial monocytes responded to activation using cytokines (IFNγ, IL-6 and IL-4) or LPS. The cells were primed for STAT1-(p=0.0039), resistant to STAT3-(p=0.0039) and had unchanged STAT6-(p=0.7344) phosphorylation compared to circulating monocytes (**figure 1G**) and displayed resistance to cytokine production of IL-1ß, IL-6, IL-8 (all p=0.0078) and TNF (p=0.0234) following activation with LPS (**figure 1H)**. Thus, synovial monocytes show signs of exhaustion and previous activation. Taken together, these data highlight a functional imbalance of monocytes in the pathogenesis; as they: (1) drive inflammation through T-cell activation, (2) are primed for STAT1- and resistant to STAT3 activation, (3) display increased clearance of apoptotic cells and (4) show resistance to cytokine production.

### Synovial fluid polarization induces a regulatory, pro-clearance phenotype and downregulates co-stimulatory capabilities in healthy monocytes

To investigate the effect of SF on the function and phenotype of monocytes, we cultured healthy monocytes overnight with SF or paired serum (as a control). Monocytes polarized with SF induced less proliferation (p<0.0001) and activation markers (p<00001) in healthy T-cells compared to serum polarized monocytes (**figure 2A and B**). Accordingly, SF-polarized monocytes expressed less CD86 and HLA (p<0.0001, **figure 2C**). In addition, SF-polarization resulted in an increased uptake of apoptotic neutrophils (p<0.0001, **figure 2D**) as well as elevated expression of markers related to clearance (CD16 and MerTK, p<0.0001, **figure 2E**). We have previously shown that synovial monocytes have reduced capacity to phagocytose and produce ROS (7). Here, we instead observed that SF-polarization induced an increase in phagocytosis (p<0.0001, **figure 2F**) and ROS production over time in healthy monocytes (**figure 2G**). Finally, monocytes polarized with SF produce less pro-inflammatory cytokines (IL-1ß, IL-8, TNF (p=0.0002) and IL-6 (p=0.0007) upon LPS activation **figure 2H**). These results suggest that SF from oJIA patients downregulates antigen presentation capabilities and induce a regulatory phenotype in healthy monocytes, mimicking some of the features of the patients’ synovial monocytes.

**Figure 2.**
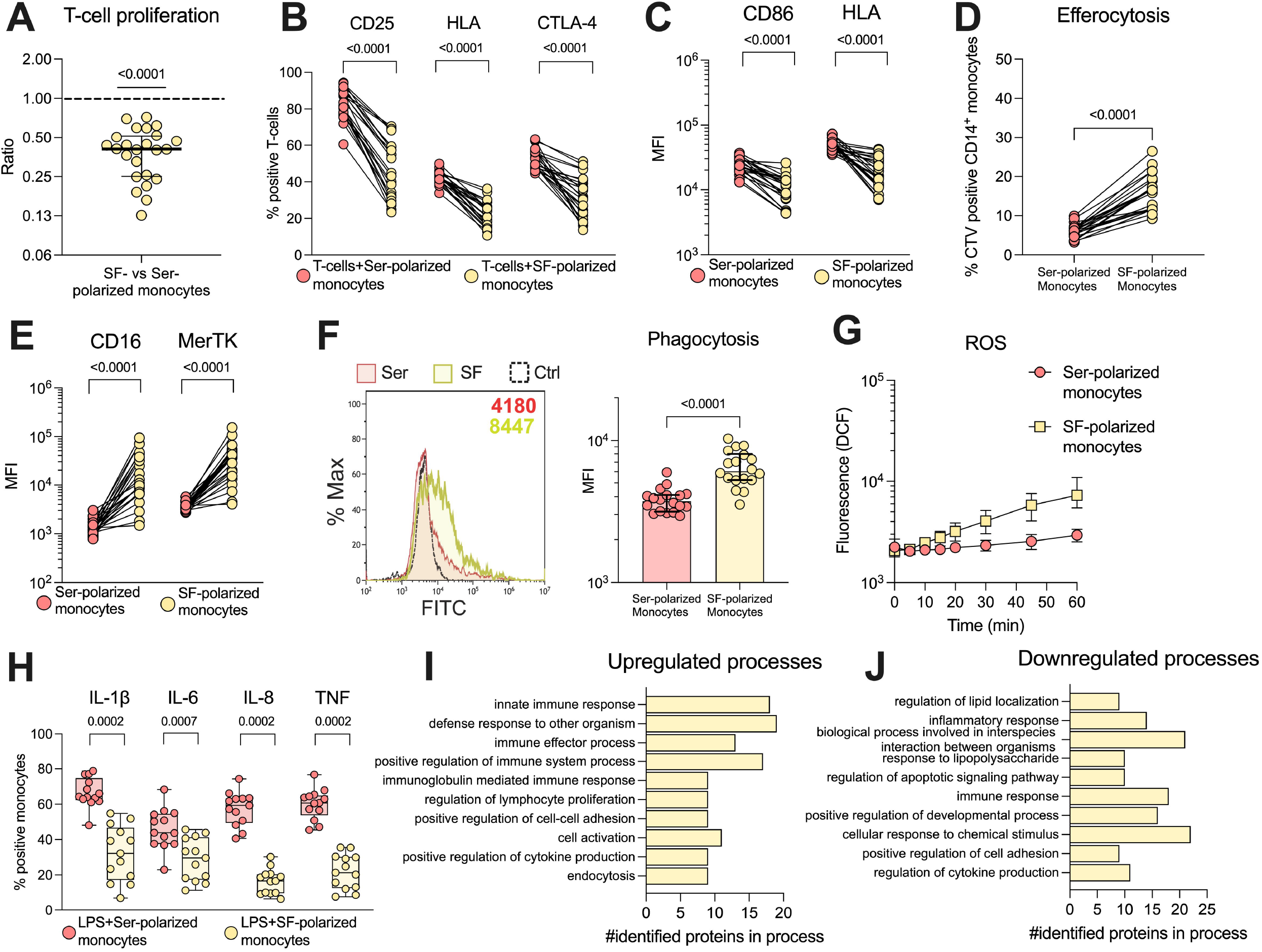
Synovial fluid induces a regulatory phenotype in healthy monocytes and downregulates co-stimulatory capabilities. Monocytes were isolated from healthy controls and polarized with 20% serum (Ser) or 20% synovial fluid (SF) overnight. Polarized monocytes were detached and co-cultured with healthy CD3-activated T-cells (1:10 monocytes to T-cells) for 72hrs and analyzed by flow cytometry for (**A**) proliferation (n=24, data is presented as the ratio of proliferation induced by SF polarized monocytes vs serum polarized monocytes) and (**B**) activation markers on T-cells (n=24). (**C**) Shows expression of CD86 and HLA in polarized monocytes (n=24). (**D**) Monocytes were incubated with CTV-stained apoptotic neutrophils for 3hrs and analyzed for uptake of these cells. Monocytes were defined as CD14^+^CD66b^-^, and the percent CTV positive monocytes was analyzed (n=21). (**E**) Displays the expression of CD16 and MerTK in polarized monocytes. (**F**) Phagocytosis of polarized monocytes was assessed following 30min incubation with opsonized FITC labeled beads (n=18). The control received no beads. (**G**) ROS production of polarized monocytes at different time points, assessed using H_2_DCFDA staining (n=24, median with interquartile range). (**H**) Cytokine production of polarized monocytes was studied intracellularly following pre-incubation with golgiplug and LPS activation for 4 hrs (n=13). To study changes at the protein level, healthy monocytes (n=3) were polarized overnight with a pool of 20% SF or paired serum and analyzed by liquid-chromatography mass spectrometry. Changes in protein expression were analyzed through biological process enrichment, and the top 10 (**I**) upregulated and (**J**) downregulated processes are presented, sorted on p-value. Statistical analyses were performed using Wilcoxon matched pairs signed rank test. Lines at median with IQR. *MFI-Median fluorescence intensity, IL-Interleukin, CTV-Cell Trace Violet, LPS-Lipopolysaccharide, ROS-Reactive oxygen species, TNF-Tumor necrosis factor*.

### Biological processes involved in immune- and regulatory processes are upregulated by synovial fluid

To explore the effect of SF at a broader scale, we analysed healthy monocytes stimulated with SF by liquid-chromatography mass spectrometry. We identified 62 upregulated- and 66 downregulated proteins (see **supplementary figure 4** for details on selection). A complete list of up- and downregulated proteins can be found in **supplementary table 1**. To understand the involvement of these proteins, we performed enrichment analysis of biological processes using gene ontology. The top 10 enriched biological processes of regulated proteins are presented in **figure 2I and J**. Generally, the upregulated processes are immune- and regulatory processes, such as innate immune response, and regulation of immune effector process, lymphocyte proliferation and cell-cell adhesion. The downregulated processes are involved in lipid localization, inflammation, and regulation of apoptosis. These findings support our previous results and suggest that SF induces upregulation of immune- and regulatory processes.

### Synovial fluid predominately induces STAT3 phosphorylation through IL-6 signalling

To characterize the major signalling pathways induced by SF, we performed a broad-spectrum phosphorylation assay, using healthy monocytes and SF from n=4 patients. SF predominately induced phosphorylation of STAT3 (pSTAT3) and, to a lesser degree, p53, compared to serum (**figure 3A**). In addition to the proteins involved in the broad-spectrum phosphorylation assay, we were interested in NFκB, a downstream signalling transducer of TNF. NFκB phosphorylation was investigated separately by flow cytometry, where we could not detect an increased phosphorylation in cells stimulated with SF compared to paired serum (p=0.3748, **figure 3B**). As STAT3 is involved in immune function, we continued to investigate this factor and could confirm the findings that SF induces pSTAT3 in a larger sample population (n=24, p<0.0001, **figure 3C**). Next, we reanalysed the IL-6 levels in SF, as published previously (7), and observed a strong correlation between IL-6 and pSTAT3 (n=31, spearman r=0.819, p<0.0001, **figure 3D**). To confirm that IL-6 is responsible for pSTAT3, we used two inhibitors of the JAK/STAT signalling pathway: the anti-IL-6-R antibody tocilizumab and the small molecule inhibitor of JAK, tofacitinib. Both tocilizumab and tofacitinib fully inhibited pSTAT3 (p<0.0001) and pSTAT1 (p<0.0001), both of which are induced by IL-6 (**figure 3E and F**). Taken together, these data suggest that SF predominately induces monocyte activation via STAT1/3 phosphorylation in healthy monocytes through an IL-6 driven mechanism.

**Figure 3.**
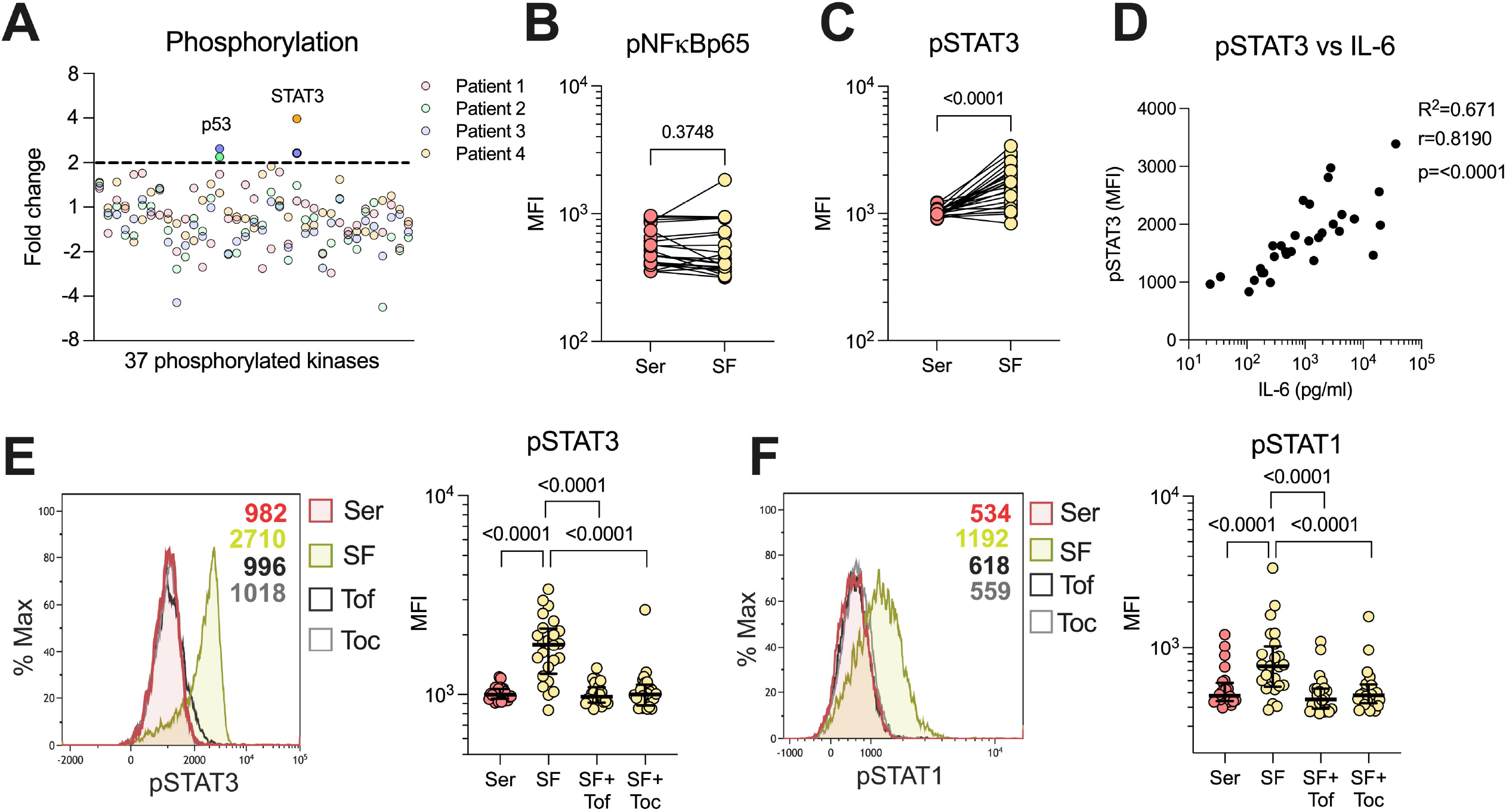
Synovial fluid predominately induces STAT3 phosphorylation through IL-6. (**A**) Broad spectrum phosphorylation array of 37 kinases following 20% synovial fluid (SF) stimulation of healthy monocytes compared to serum (n=4). (**B**) Analysis of NFkBp65 phosphorylation with SF vs serum (Ser) stimulation by flow cytometry (n=24). (**C**) Analysis of STAT3 phosphorylation with SF (n=24) by flow cytometry. (**D**) Shows correlation between levels of STAT3 phosphorylation in monocytes versus IL-6 in the SF (n=33, Spearman correlation). (**E**) SF stimulation and STAT3 phosphorylation analysis, with pre-treatment of monocytes with tofacitinib (1µM) or tocilizumab (100ng/ml) and (**F**) STAT1 phosphorylation (n=24). Data is presented as median with IQR. *MFI-Median fluorescence intensity, IL-Interleukin, STAT-Signal transducer and activator of transcription, Tof-Tofacitinib, Toc-Tocilizumab*.

### Blocking of IL-6/JAK/STAT signalling inhibits several aspects of the synovial fluid polarized phenotype

To investigate the role of IL-6/JAK/STAT on the functional alterations observed in the patients’ monocytes, we pre-treated healthy monocytes with tocilizumab or tofacitinib, followed by polarization with SF. Tocilizumab (p=0.0056) and tofacitinib (p=0.0192) restored the decreased T-cell proliferation to some extent, and had mild effects on the T-cell expressed activation markers (**figure 4A and B**). Yet, the CD86 and HLA expression was restored upon inhibition of IL-6/JAK/STAT (p<0.0001, **figure 4C**). Neither drug had any substantial effect on the increased efferocytosis (p=0.5449 and p=0.3843, **figure 4D**), but efficiently restored the CD16 and MerTK expression (all p<0.0001, **figure 4E**). Both drugs also inhibited the increased phagocytosis (p<0.0001, **figure 4F**), and tofacitinib partly inhibited the increased ROS production (p<0.0001, **figure 4G**). Finally, the SF induced resistance to LPS activated cytokine production was inhibited by both drugs (all p=0.0006, **figure 4H**). These results suggest that the IL-6/JAK/STAT pathway is responsible for several of the regulatory features observed in synovial monocytes and highlight potential targets of anti-IL-6/JAK/STAT therapy.

**Figure 4.**
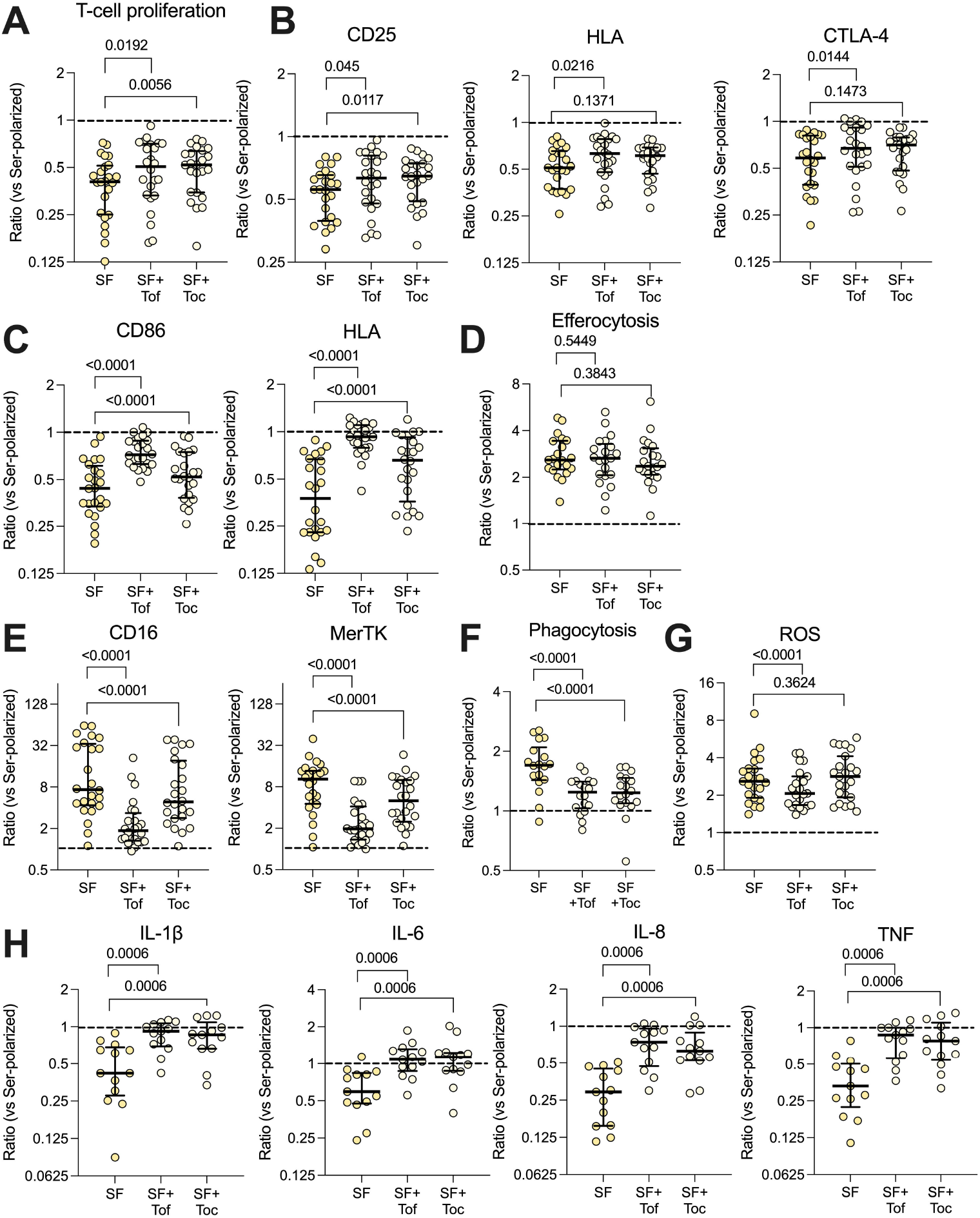
The IL-6/JAK/STAT axis is responsible for several of the phenotypical effects induced by synovial fluid. Healthy monocytes were pre-treated with or without tofacitinib (Tof, 1µM) or tocilizumab (Toc, 100ng/ml) and then polarized with 20% serum (Ser) or 20% synovial fluid (SF). Polarized monocytes were incubated with healthy T-cells for 72hrs and assessed for (**A**) proliferation of T-cells (n=24) and (**B**) activation markers on the T-cells. (**C**) Shows expression of CD86 and HLA in polarized monocytes (n=24). (**D**) Efferocytosis of CTV-stained apoptotic neutrophils by CD14^+^CD66b^-^ monocytes (n=21). (**E**) CD16 and MerTK expression in polarized monocytes. (**F**) Phagocytosis of opsonized FITC labeled beads (n=18). (**G**) Shows ROS production after 1hr of incubation (n=24). (**H**) Intracellular cytokine production following pre-incubation with golgiplug and LPS activation for 4 hrs (n=13). Data represents the ratio vs serum-polarized monocytes, and the dotted lines represents a ratio of 1:1. Full lines at median with IQR, Wilcoxon matched pairs signed rank test. *ROS-reactive oxygen species, CTV-Cell Trace Violet, LPS-Lipopolysaccharide, MFI-Median fluorescence intensity, IL-Interleukin, STAT-Signal transducer and activator of transcription, Tof-Tofacitinib, Toc-Tocilizumab, LPS-Lipopolysaccharide, TNF-Tumor necrosis factor*.

### Cell-cell interactions induce increased antigen presentation capabilities in synovial fluid polarized monocytes

SF polarization of healthy monocytes does not replicate the increased co-stimulatory capabilities observed in the patients’ synovial monocytes; nor does it induce a decrease in ROS production and phagocytosis, as previously observed (7). We have recently shown that migration through a transwell system influences the effector functions of neutrophils (8). To determine how the patient’s synovial monocytes may acquire the remaining features, we investigated the effect of migration through an artificial synovial membrane towards SF on healthy monocytes (**supplementary figure 5A**). We show that migration induced some, but not all, of the features of the patients’ monocytes, such as increased CD86 expression and increased T-cell proliferation (**supplementary figure 5B-F**). We theorized that the monocytes may instead require prolonged cell-cell contact to induce the remaining features. Thus, we studied the effect of co-culture between monocytes and FLS in SF-polarizing environment (**figure 5A)**. Co-culture resulted in an increased ability of the monocytes to induce T-cell activation (p<0.0001, **figure 5B and C**). Additionally, these monocytes had increased expression of both CD86 and HLA (p<0.0001, **figure 5D**) as well as reduced ROS production (p<0.0001, **figure 5E**) and phagocytosis (p=0.0005, **figure 5F**). Finally, tofacitinib and tocilizumab did not inhibit the increased expression of CD86, HLA or T-cell proliferation, but rather trended to increase it further, in both migrated and co-cultured monocytes (**supplementary figure 6**). Thus, co-culture with FLS induces the remaining features in healthy monocytes that are observed in the patients’ synovial monocytes, such as increased co-stimulatory capabilities. These data suggest that cell-cell contact promotes inflammatory monocytes and highlight the monocytes’ potential to drive adaptive immune responses.

**Figure 5.**
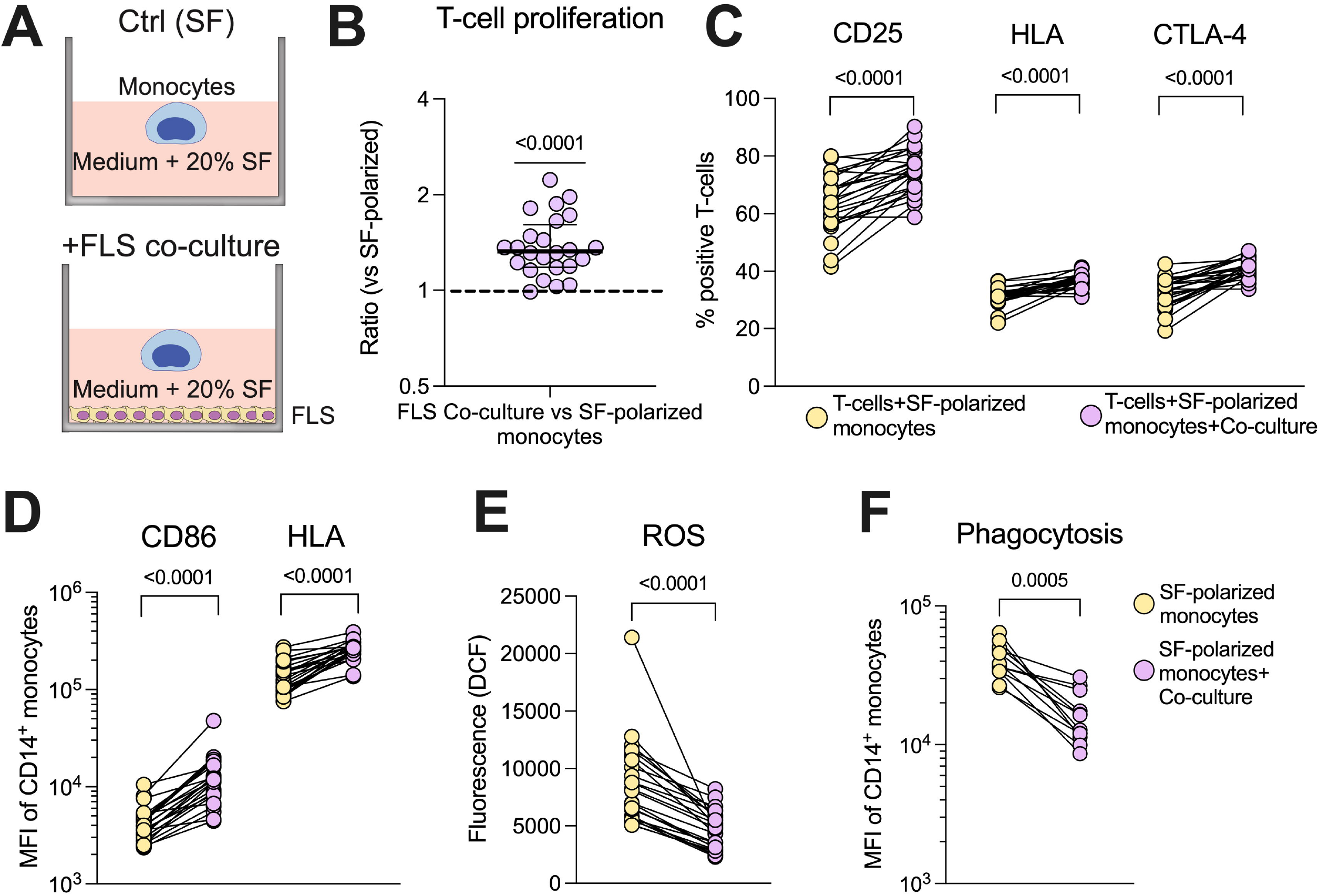
Increased co-stimulatory capabilities of synovial monocytes are induced *in vitro* in healthy monocytes through co-culture with fibroblast-like synoviocytes. (**A**) Experimental setup of the co-culture assay. (**B**) Mono-culture of monocytes (SF) and co-culture with FLS (Co-culture) monocytes were detached following overnight culture and seeded with T-cells (1:10 monocytes to T-cells) for 72hrs, followed by analysis of proliferation (displayed as ratio of percent proliferation between FLS co-culture vs mono-culture of monocytes) and (**C**) expression of activation markers in T-cells (n=24). (**D**) Shows changes in surface expression of CD86 and HLA. (**E**) Displays ROS production after 1hr incubation following H_2_DCFDA staining (n=24) and (**F**) phagocytosis of opsonized FITC labeled beads (n=12). Wilcoxon matched pairs signed rank test. Lines at median. *FLS-Fibroblast-like synoviocytes, ROS-Reactive oxygen species, MFI-Median fluorescence intensity, SF-Synovial fluid*.

### The IL-6/JAK/STAT axis activation in monocytes is reflected in circulating markers of inflammation

Given the effect of IL-6/JAK/STAT on the monocyte phenotype, we speculated that the synovial IL-6/JAK/STAT driven inflammation is reflected in the blood. Using hierarchical clustering based on three parameters, synovial IL-6, pSTAT1- and pSTAT3, we generated two different groups of low-vs high synovial IL-6 driven inflammation. We subsequently depicted Z-score pattern of several prominent markers of inflammation in plasma (IFNα2a, IL-6, SAA, CRP, IL-8, and TNF) to highlight any differences between the two groups (**figure 6A**). We next used a random forest model using the plasma markers to predict the groups. IFNα2a and IL-6 were ranked the highest in the variable importance scores (**figure 6B**). Thereafter, we analysed the differential expression of the circulating markers between the two groups, and we observed that most markers were significantly elevated in the group with prominent synovial IL-6/JAK/STAT driven inflammation (**figure 6C**). Clinically, there were no major differences between the two groups, although patients in group two tended to be younger and had an overrepresentation of males (**supplementary figure 7**). These results suggest that the magnitude of synovial inflammation related to the IL-6/JAK/STAT axis is reflected in the circulation, and the possibility to identify patients more relevant for anti-IL-6/JAK/STAT therapy. Notably, this is apparent in several different circulating markers.

**Figure 6.**
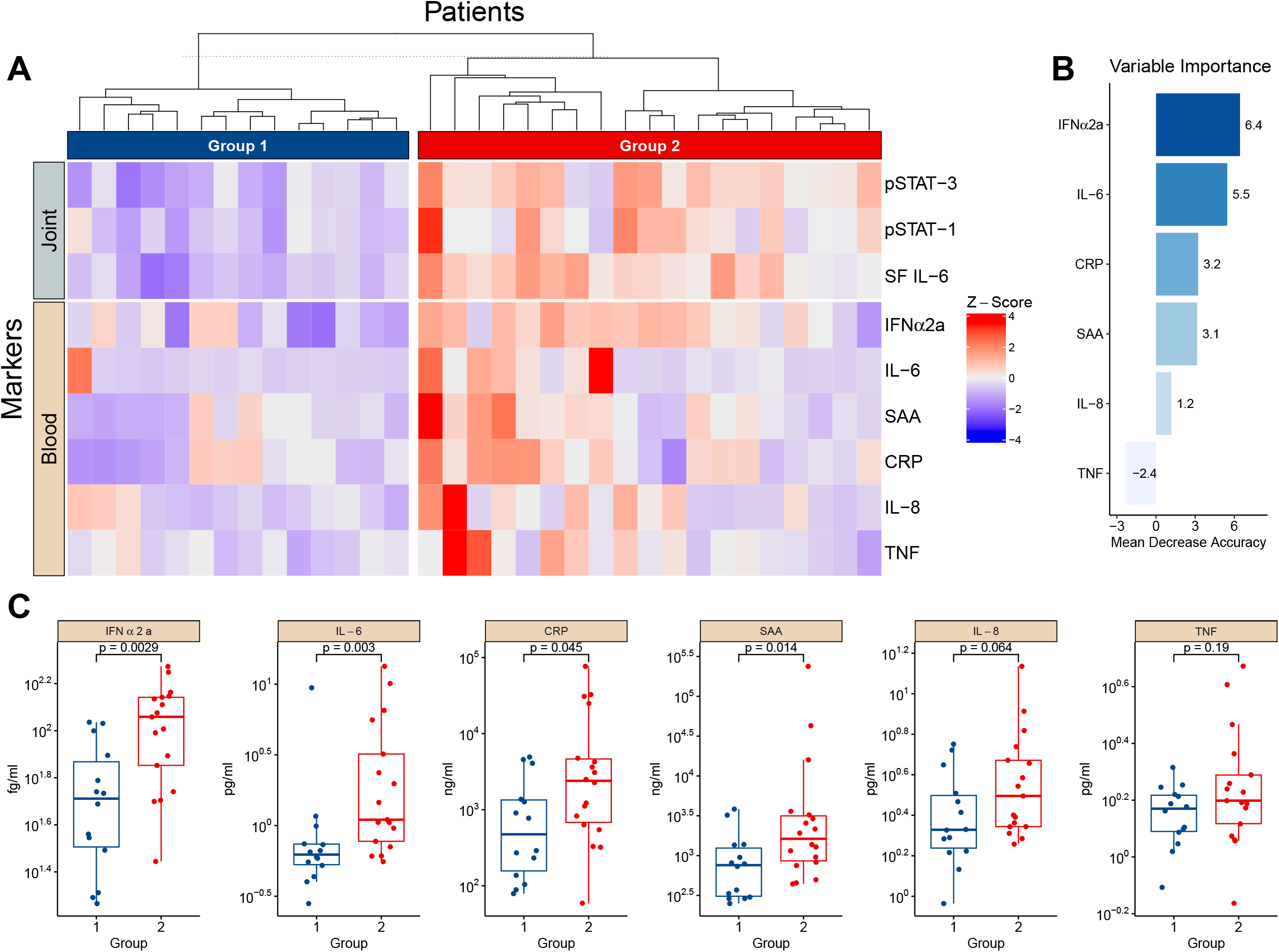
The magnitude of the synovial IL-6/JAK/STAT axis is reflected in several circulating markers of inflammation. (**A**) Heatmap showing hierarchical clustering into two groups based on three parameters, synovial (joint) IL-6 levels and monocyte pSTAT1- and pSTAT3 activation. It further shows the distribution of circulating (blood) markers of inflammation measured in plasma. (**B**) Shows the importance score of the circulating markers in predicting the two groups based on a random forest predictive model. (**C**) Displays the statistical difference between group one and two regarding the levels of circulating markers. Statistical analyses between groups were performed using Mann-Whitney U test. *SF-Synovial fluid, IFN-Interferon, CRP-C-reactive protein, SAA-Serum amyloid A, TNF-Tumor necrosis factor*.

## Discussion

Monocytes are important cells of the innate immune system with the potential to drive inflammation. Here, we show that synovial monocytes from patients with untreated oligoarticular juvenile idiopathic arthritis (oJIA) have altered functions with implications in both inflammatory- and regulatory processes. The regulatory aspects can be replicated *in vitro* through polarization of healthy monocytes with synovial fluid (SF), which is mainly driven through an IL-6/JAK/STAT mechanism. The magnitude of IL-6/JAK/STAT activation is also reflected in circulating markers of inflammation. The co-stimulatory aspects are induced by cell-cell interactions (i.e., co-culture with fibroblast-like synoviocytes (FLS)). These data provide insight into the pathogenesis of oJIA and suggest that monocytes drive chronic inflammation, e.g., through prolongating T-cell responses. Interestingly, the magnitude of IL-6/JAK/STAT activation is reflected in circulating markers of inflammation, implying that we could identify a group of patients likely to benefit from IL-6/JAK/STAT inhibition, suggesting use in personalized medicine for the treatment of oJIA.

In adult arthritis, monocytes are found in the inflamed synovium, expressing markers of activation, producing cytokines, and can activate other immune cells, such as T-cells (11, 17, 25). Less is known of monocytes in JIA. We have previously shown that synovial monocytes in oJIA are polarized, displaying markers of activation, such as co-stimulatory molecules (7). A recent study found epigenetic alterations related to an activated phenotype in synovial monocytes from oJIA patients (10). In addition, synovial T-cells display an activated phenotype and produce several pro-inflammatory cytokines, such as IL-17 (26-28). Here, we confirmed that synovial monocytes express high levels of co-stimulatory markers and further showed that the monocytes induce proliferation and activation of T-cells. We have recently shown that synovial neutrophils lose their capacity to suppress T-cell proliferation, and that healthy neutrophils lose this ability upon migration (8). Thus, an increase in activation of T-cells by monocytes, and a loss of suppressive capability by neutrophils may explain the expansion and activation of T-cells within the joint of patients with oJIA.

Notably, this is not an effect of the environment on monocytes alone, as SF polarization reduced the ability of monocytes to activate T-cells. Indeed, this reduced ability to induce T-cell activation is supported by our proteomics data, where regulation of lymphocyte proliferation was one of the top upregulated processes. This process included proteins such as VSIG4 and GPNMB, which are described to negatively regulate T-cell proliferation (29, 30). However, co-culture with healthy FLS, or migration to a certain extent, resulted in an increased ability of the monocytes to induce T-cell activation compared to SF polarization alone, suggesting a role of cell-cell interactions in driving the synovial monocyte phenotype. A link between monocytes and FLS is well established; monocytes can induce cytokine production in synoviocytes, such as IL-6, whilst synoviocytes produce factors such as GM-CSF, that in turn can induce HLA and increase viability in monocytes (31-33). A direct cell-cell contact may be important in the interaction of these cells and can induce factors such as MIP-1α (34). Finally, synoviocytes in JIA upregulate VCAM-1, which may facilitate leukocyte recruitment, attachment, and activation (35, 36). We are going to pursue the interaction between JIA FLS and monocytes in oJIA in future studies.

Synovial monocytes also displayed signs of a regulatory phenotype; features which could all be replicated *in vitro* following SF polarization, driven partly by IL-6/JAK/STAT signalling. IL-6/JAK/STAT signalling appears to have a central role in arthritis, where it is believed to drive recruitment, inflammation, and phenotypical changes in T-cells, B-cells and FLS (37-39). Indeed, STAT3 knockout is protective in a mouse model of arthritis; and tofacitinib treatment reduced synovial matrix metalloproteinases, chemokines, and phosphorylation of STAT3, which in turn correlated with clinical responses in rheumatoid arthritis (40, 41). A role of IL-6 in oJIA has been implied decades ago (42). In addition, a recent study investigating chromatin data in monocytes in JIA patients found that IL-6 binding was one of the top enriched processes (9). *Peeters. et. al*. found that inhibition of JAK/STAT signalling blocked an inflammatory phenotype at the epigenetic level in monocytes from oJIA patients (10). Interestingly, our data showed that the magnitude of IL-6/JAK/STAT activation in monocytes was reflected in the levels of several circulating cytokines. By clustering the patients into two groups based on the synovial IL-6/JAK/STAT axis, we observed that patients with high IL-6/JAK/STAT involvement also had significantly higher levels of several circulating cytokines, with IFNα2a and IL-6 being the most important variables in predicting the two groups. Notably, the impact of TNF was minor. Previous studies have also investigated the relationship between circulating- and synovial inflammatory markers (43, 44). These studies also suggest potential differences, both between and within JIA subgroups. Even though oJIA is not traditionally associated with systemic inflammation, our results further support that, following future studies, synovial or circulating markers could be important in treatment prediction, and the division of patients with a more IL-6 prominent disease.

Yet, in line with our results, the effect of IL-6/JAK/STAT signalling on monocytes/macrophages is believed to be primarily regulatory/anti-inflammatory (45-47). An impaired efferocytosis, resulting in accumulation of cellular debris and increased autoantigenic burden, can further autoimmune reactions (48). However, we did instead observe an increased efferocytosis of synovial monocytes and SF-polarized healthy monocytes. Additionally, SF polarization also induced an increase in phagocytosis, whilst synovial monocytes in oJIA have a decreased ability to phagocytose (7). Though, this reduced phagocytosis could be replicated following co-culture with FLS. Thus, synovial monocytes might have impaired clearance of certain pathways, whilst remaining functional in others. Furthermore, we observed a reduction of pro-inflammatory cytokine production following activation with LPS, suggesting resistance to a pro-inflammatory response. This was dependent on IL-6 signalling, which has previously been described to impair LPS induced cytokine production (49). Given the presence of endogenous toll-like receptor (TLR) ligands in the joint and the differential expression of TLRs on synovial monocytes, IL-6 could play a role in regulating TLR induced cytokine production in the joint (11, 22, 49). Importantly, regulatory monocytes likely still contribute to chronic inflammation. For example, regulatory monocytes and macrophages are prominent in angiogenesis, a process believed to be unfavourable in arthritis (19, 20). CD163 expression, a classical marker of regulatory macrophages, correlates with inflammation in spondylarthritis (21). In addition, regulatory monocytes and macrophages are potent inflammatory cytokine producers in arthritis (22). Taken together, our data suggests that SF mainly induces a regulatory phenotype in monocytes, but this phenotype may not be advantageous in chronic disease, and we suggest that it would be beneficial to restore synovial homeostasis blocking IL-6/JAK/STAT, rather than favouring anti-inflammation. In addition, treatment of the patients’ monocytes with JAK/STAT inhibitors reverses epigenetic changes, further supporting blockage of this pathway to restore homeostasis (10). Finally, blocking of monocytes from entering the joint, and subsequent cell-cell interactions, might represent another crucial mechanism.

There are some limitations to this study. There are several aspects not investigated in this study that may influence monocyte function *in vivo*, such as time spent in the joint, hypoxia, and biomechanical factors (e.g., tissue stiffness). Even though the phosphorylation assay contained several of the common kinases involved in monocyte activation, there are still numerous pathways not included, that could also contribute to the resulting phenotype. Indeed, blocking IL-6/JAK/STAT did not normalize all features induced by SF, suggesting a role of other pathways/factors in SF. The statistical changes in the proteomics analysis should be interpreted carefully, as we analysed 3 donors, the statistical power is low and false discovery rate could not be accounted for. Thus, it should be considered supportive rather than conclusive. Finally, the T-cell interaction assays used in this study involves pan TCR activation using an anti-CD3 antibody. Thus, the extent to how monocytes contribute to inflammation by antigen presentation through MHC, and which antigens that are of importance, remains to be determined.

In conclusion, we show that synovial monocytes contribute to the pathogenesis of oJIA by driving chronic inflammation. Their phenotype could be replicated *in vitro* using SF, which induced regulatory aspects through IL-6/JAK/STAT signalling, and co-culture with FLS, which induced co-stimulatory properties. In addition, the synovial IL-6/JAK/STAT axis was reflected in circulating markers, suggesting a group of patients more likely to benefit from targeting the IL-6/JAK/STAT axis to restore synovial homeostasis.

## Data Availability

All data produced in the present study are available upon reasonable request to the authors. The raw proteome data is available via ProteomeXchange with identifier PXD033983.

http://www.proteomexchange.org/

## Acknowledgements and affiliations

We wish to thank Julia Westerlund for T cell protocols and reagents. We also thank Charlotte Welinder and the Center for Translational Proteomics at Lund University and Region Skåne, for assistance with proteomics analysis. Some of the findings of this work have previously been reported at Paediatric Rheumatology European Society (PReS) conference in 2022 (50). This study was supported by grants from the Royal Physiographic Society of Lund (to TS). RK is funded by grants from the Swedish League against Rheumatism, Greta and Johan Kock’s Foundation, the Anna-Greta Crafoord Foundation, the Crafoord Foundation, Alfred Österlunds Foundation, Magnus Bergvall Foundation, King Gustaf V’s 80-year foundation, The Knut and Alice Wallenberg foundation, the Medical Faculty at Lund University and Region Skåne. The funders had no role in the concept, design, or interpretation of data.

## Author contributions

TS conceptualized the study, designed experiments, analysed, and interpreted data, and wrote the manuscript. AM, SAB, and ER performed experiments, interpreted data, and reviewed and revised the manuscript. EB, PK, AD, and BM collected clinical data and samples, interpreted data, and reviewed and revised the manuscript. SMN analysed and interpreted statistical data and reviewed and revised the manuscript. AB and FK interpreted data and reviewed and revised the manuscript. RK conceptualized the study, collected clinical data and samples, interpreted data, and wrote the manuscript. All authors have read and approved the final manuscript.

## Notes

### Competing Interest Statement

The authors have declared no competing interest.

### Funding Statement

RK is funded by grants from the Swedish League against Rheumatism, Greta and Johan Kocks Foundation, the Anna-Greta Crafoord Foundation, the Crafoord Foundation, Alfred Osterlunds Foundation, Magnus Bergvall Foundation, King Gustaf Vs 80-year foundation, The Knut and Alice Wallenberg foundation, the Medical Faculty at Lund University and Region Skane.
The funders had no role in the concept, design, or interpretation of data.

### Author Declarations

The Regional Ethical Review Board for southern Sweden (2016/128) gave ethical approval for this work.

