## Supplementary Materials and Methods for "Synovial Monocytes Drive the Pathogenesis in Oligoarticular Juvenile Idiopathic Arthritis via IL-6/JAK/STAT Signalling and Cell-Cell Interactions"

**­Sample Preparation**

EDTA blood was used as control for analysis of surface markers (see below). The serum blood was allowed to clot for 1hr at room temperature followed by centrifugation at 1900g, 10min. Next, the serum was aliquoted and stored at -80°C until use. The heparin blood was used as a control for functional assays and monocyte isolation (described below).

A proportion of synovial fluid (SF) was used for functional studies and the rest was centrifuged at 500g, 10min. The SF was collected and centrifuged a second time at 800g, 10min to generate cell-free SF. Then, it was frozen at -80°C in aliquots until use. The remaining synovial cell fraction was washed once with PBS and subsequently resuspended in PBS with 0.5% BSA. Peripheral blood mononuclear cells (PBMCs), from heparinized blood, and synovial fluid mononuclear cells (SFMCs), from the synovial cell fraction, were isolated through density gradient centrifugation (Lymphoprep, Axis-Shield) at 620g, 20min with low break. The PBMCs and SFMCs were washed twice with PBS. Monocytes were further isolated from PBMCs and SFMCs using CD14^+^ magnetic bead separation (Miltenyi) as per the manufacturer’s instructions. The purified monocytes were counted and then used in subsequent assays described below.

**Mesoscale cytokine measurement**

CRP and SAA were measured in plasma (diluted 1/5000) using the vascular injury panel 2 (Mesoscale Diagnostics) according to the manufacturer’s instructions. IL-1, IL-6, IL-8 and TNF were analysed in plasma (diluted 1/2) and SF (diluted 1/25) using the pro-inflammatory panel 2 (4-plex). Finally, IFNα2a was measured in plasma and SF (both neat) using the S-PLEX human IFNα2a kit. Data was processed and concentrations were calculated using the Discovery Workbench software (Mesoscale, version 4.0).

**Monocyte Isolation and Polarization *in vitro***

Monocytes were isolated from freshly isolated PBMCs, as described above, from healthy controls upon informed consent. Monocytes were next cultured overnight in 96-well plates (Falcon) at 1x10^6^cells/ml in RPMI-1640 medium with 2.05mM L-glutamine supplemented with either 20% patient serum or 20% paired SF to generate *in vitro* polarized monocytes. Each experimental section was performed using monocytes from at least two different donors unless otherwise stated. For blocking assays, monocytes were pre-incubated for 25min at 37°C with 1µM tofacitinib or 100ng/ml tocilizumab before the addition of SF. The concentrations of tofacitinib and tocilizumab used are in line with previous reports (1, 2). Accordingly, we confirmed that the effect of these drugs on cell viability were minor, using annexin V / PI exclusion of apoptotic and dead cells (**supplementary figure 1**).

**Surface Marker Analysis**

For patients, synovial cells in PBS with 0.5% BSA at 2x10^6^cells/ml or 100µl of EDTA blood were incubated with two antibody mixes. First mix: anti-CD3 (clone UCHT-1, BV786, 1:50, BD), CD19 (clone HIB-19, Bv786, 1:125, BD), CD56 (clone NCAM 16.2, BV786, 1:125, BD), CD14 (clone 63D3, 1:50, Biolegend), CD66b (clone G10F5, alexa fluor 647, 1:50, BD), CD16 (clone 3G8, PerCP Cy5.5, 1:50, BD), MerTK (clone 590H11G1E3, PE, 1:100, Biolegend) and CD86 (clone FUN-1, BV650, 1:100, BD). The second mix contained the lineage antibodies (CD3, CD19, CD56, CD66b and CD14) and anti-HLA-DR, DP, DQ (clone Tü39, PerCP Cy5.5, 1:100, Biolegend) for 25min, RT. The cells were washed once with PBS before analysis using flow cytometry (CytoFLEX, Beckman Coulter). Gating strategy can be found in **supplementary figure 2A**.

*In vitro* polarized monocytes were detached using ice-cold PBS/1mM EDTA and gentle pipetting. Next, they were washed with PBS and stained with anti-CD16, MerTK, CD86 and HLA, all diluted 1:200, for 25min, RT. Finally, they were washed once more with PBS and analysed by flow cytometry.

**STAT Phosphorylation**

For patients, 100µl of heparinized blood or 100µl of whole SF in polypropylene FACS tubes (Falcon) were stained with (CD66b (clone G10F5, alexa fluor 700, 1:100, Biolegend), CD14 (clone HCD14, BV421, 1:100, Biolegend), CD3 (clone UCHT-1, BV510, 1:100, BD), CD19 (clone SJ25C1, BV510, 1:200, BD) and CD56 (NCAM16.2, BV510, 1:200, BD) in 100µl of PBS with 0.5% BSA. Simultaneously IFNγ (5ng/ml, R&D Systems), IL-4 (5ng/ml, R&D Systems), and IL-6 (5ng/ml, R&D Systems) were added to one set of tubes. Tubes not receiving cytokines and unstained tubes served as controls. The cells were incubated for 15min, 37°C. Next, 2ml of lyse/fix (BD) was added to each tube and incubated for another 10min, 37°C followed by centrifugation at 500g, 8min. The cells were then washed once with PBS and permeabilized as described below.

For *in vitro*, monocytes from healthy controls at 1x10^6^/ml in RPMI-1640 medium supplemented with 0.2% BSA were stimulated with 20% SF or 20% paired serum for 10min, 37°C to induce phosphorylation. In some experiments, monocytes were pre-incubated for 25min, 37°C with tofacitinib or tocilizumab (see above) before stimulation. Monocytes were subsequently fixated (CytoFix, BD) for 15min, 37°C before centrifugation.

Cells, either patient monocytes or *in vitro* polarized monocytes, were permeabilized (Perm Buffer III, BD) for 30min on ice followed by two washes with PBS. The patients’ monocytes were next stained for anti-STAT1-pY701 (clone 4a, alexa fluor 647, 1:100, BD), STAT3-pY705 (clone 4/P-STAT3, PE, 1:100, BD) and STAT6-pY641 (clone 18/p-Stat6, alexa fluor 488, 1:100, BD), whilst the *in vitro* polarized monocytes were stained with STAT1, STAT3 and NFkBp65-pS529 (clone K10-895, alexa fluor 488, 1:100, BD) in PBS supplemented with 0.5% BSA for 25min. Finally, the monocytes were washed once with PBS before analysis (CytoFLEX). Gating strategy for analysis of STAT phosphorylation in patients can be found in **supplementary figure 2B**.

**T-cell Isolation and Proliferation**

PBMCs were isolated from healthy controls using density centrifugation as described previously. CD4^+^ T-cells were isolated from the PBMC fraction using the EasySep^TM^ CD4^+^ T-cell isolation kit (Stemcell Technologies) as per the manufacturer’s instructions. The T-cells were stained with 2µM CellTrace Violet (Invitrogen). A 96-well plate (Eppendorf) was coated with anti-CD3 (1:1000, Clone OKT3, Invitrogen) for 90min. The coating solution was removed prior to use. Wells without coating served as negative controls.

Monocytes from patients or *in vitro* polarization, as described above, were counted (XN-350, Sysmex) and resuspended in RPMI-1640 supplemented with 10% fetal calf serum, 2mM L-glutamine and PenStrep. Next, monocytes and T-cells at a 1:10 ratio (monocytes:T-cells) were added to the coated plate in a total volume of 200µl. The cells were incubated for 72h, 37°C, at 5% CO_2_. The cells were detached through gentle pipetting, centrifuged, and stained with anti-CD3 (clone UCHT1, alexa fluor 700, 1:200), anti-CD25 (clone M-A251, PerCP Cy5.51:200), anti-HLA-DR (clone G46-6, APC-H71:200) and anti-CTLA-4 (clone BNI3, PE, 1:50), all from BD, for 25min, RT. Finally, the cells were washed once with PBS and analysed using flow cytometry (CytoFLEX).

**LPS activated Cytokine Production**

For patients, heparinzed blood or fresh synovial fluid were diluted 1:1 with RPMI-1640 medium in polypropylene FACS tubes. Next, 0.5µl of golgiplug (BD) was added followed or not by 10ng/ml of LPS. The cells were incubated for 4hrs, 37°C, 5% CO_2_. For the last 15min, the cells were stained with anti-CD3 (clone UCHT-1, BV510, BD, 1:125), anti-CD19 (clone HIB19, BV786, BD, 1:150), CD14 (clone 63D3, alexa fluor (AF) 700, Biolegend, 1:125). Cells were fixated and lysed with BD lyse/fix solution for 10min, 37°C and subsequently washed two times with PBS. Permeabilization was performed by 10min incubation with 1ml of BD wash/perm (BD) followed by centrifugation and staining with anti-IL1ß (clone JK1B-1, AF647, Biolegend), IL-6 (cloneMQ2-13A5, PE/Cy7, Biolegend), IL-8 (clone E8N1, AF488, Biolegend) and TNF (cloneMAb11, BV650, BD) all diluted 1:50, for 25min, RT. The tubes were washed a final time with PBS before analysis using flow cytometry (CytoFLEX). Cells not receiving golgiplug or LPS were used to set the gates.

Healthy monocytes were resuspended in RPMI-1640 medium and seeded at 1x10^6^cells/ml with 20% serum or 20% paired SF in a 96-well plate. 0.5µl of golgiplug was added to each well followed by activation or not with 1ng/ml of LPS. The cells were incubated for 4hrs at 37°C, 5% CO_2_. Next, the cells were detached with PBS/1mM EDTA and gentle pipetting and washed once with PBS. They were subsequently fixated and permeabilized (CytoFix, BD) for 20min, 4°C. Thereafter, the monocytes were washed once with BD wash/perm and stained with the anti-IL-1ß, IL-6, IL-8 and TNF as above, all diluted 1:100 for 25min, RT. Finally, the cells were washed a final time and analysed by flow cytometry (CytoFLEX). Cells not receiving golgiplug or LPS were used to set the gates. Gating strategy can be found in **supplementary figure 2C**.

**Efferocytosis**

Neutrophils from heparinized blood of healthy donors were isolated through density centrifugation (Lymphoprep) as described above, followed by sedimentation of red blood cells for 20min using 1.5% Dextran T500 (Pharmacosmos) in saline. The remaining cells were transferred to a new tube and washed once with PBS. Residual red blood cells were lysed with sterile H_2_O for 25 seconds before restoration of isotonicity. 10^7^ neutrophils were resuspended in PBS and stained with 2µM of CellTrace Violet (CTV, Invitrogen) for 20min, 37°C. Extracellular dye was quenched by addition of RPMI-1640 medium supplemented with 10% FCS for 5min before centrifugation. The neutrophils were resuspended in serum-poor medium (RPMI-1640 medium supplemented with 1% normal human serum (Sigma-Aldrich)) at 5x10^6^ cells/ml and cultured for 24hrs at 37°C, 5% CO_2_ to induce apoptosis. Next day, the neutrophils were filtered through a cell-strainer cap (Falcon), centrifuged, and resuspended at 1x10^6^ cells/ml in RPMI-1640 supplemented with 10% of the neutrophil donor’s serum. Apoptosis was confirmed for each experiment through Annexin V staining, and monocytes were defined as CD14^+^CD66b^-^ to exclude bound, but not internalized, neutrophils (**supplementary figure 3A**).

For *in vitro* polarized monocytes (see above), medium was replaced with medium containing 1x10^5^ neutrophils, and the plate was incubated for 3hrs, 37°C, 5% CO_2_. For patients, 1x10^5^ freshly isolated monocytes, from either blood or SF, were resuspended in medium containing 1x10^5^ neutrophils, plated, and incubated as above. The cells were detached with cold PBS/1mM EDTA and gentle pipetting. Finally, the cells were washed once with PBS and stained with anti-CD14 (clone 63D3, Alexa Fluor 700, 1:200, Biolegend) and anti-CD66b (clone G10F5, FITC, 1:50, BD) for 25min, RT, followed by a final wash before analysis by flow cytometry (CytoFLEX). Monocytes were defined as CD14^+^CD66b^-^ to exclude monocytes that bound, but didn’t internalize, neutrophils (**supplementary figure 3B**). Percentage of CTV^+^ monocytes were used for analysis, and monocytes not receiving neutrophils were used to set the gates.

**Phagocytosis**

Phagocytosis was assessed using a bead-based phagocytosis assay (Cayman). Briefly, *in vitro* polarized monocytes’ medium was replaced with RPMI-1640 supplemented with 10% of the monocyte donor’s serum and diluted FITC-labelled opsonized beads (final dilution: 1/500). Binding of the beads, rather than phagocytosis, to the monocytes was assessed through incubation on ice (data not shown). Phagocytosis was performed for 30min, 37°C. Cells were detached with ice-cold PBS/1mM EDTA and surface bound beads were quenched with a 2min incubation with trypan blue on ice. Cells were subsequently washed twice with PBS before analysis by flow cytometry (CytoFLEX). The gates were set using cells that did not receive beads.

**ROS**

*In vitro* polarized monocytes were detached using PBS/1mM EDTA. The cells were washed with PBS and resuspended in 100µl RPMI-1640 supplemented with 5% NHS (Sigma-Aldrich). The cells were transferred to a black 96-well plate (Thermo Fisher) and placed in a 37°C pre-heated plate reader (VICTOR^3^, 1420 Multilabel Counter, PerkinElmer Life Sciences) for 10min. Next, 10µM H_2_DCFDA was added to each well and the plate was analysed at different time points up to 1hr of incubation. The plate was read at 485/535nm and analysed using the Wallac 1420 software (version 3.0, PerkinElmer Life Sciences) and excel. Unstained serum- and synovial fluid polarized monocytes served as controls.

**Phosphorylation Profiler Array**

From one healthy donor, 7x10^6^ isolated monocytes were exposed to 20% SF from n=4 patients with oligoarticular JIA or 20% of the monocyte donor’s serum (as a control) for 10min at 37°C. Next, cells were washed once with ice-cold PBS. Cells were then lysed, and a membrane-based 37 kinase phosphorylation array (R&D Systems) was performed according to the manufacturer’s instructions. The array was analysed by a ChemiDoc XRS+ (BioRad) and the ImageLab Software 5.1 Beta. The background was subtracted from the intensity of each dot, duplicates were averaged and the fold change of the SF- vs serum- samples was calculated.

**Liquid-Chromatography Mass Spectrometry**

Healthy monocytes from three donors were isolated using the pan monocyte isolation kit (Miltenyi) according to the manufacturer’s instructions and polarized overnight as described above with a serum pool (control) or SF pool from 6 oJIA patients. All conditions were performed in triplicates. Monocytes were detached using ice-cold PBS/1mM EDTA and gentle pipetting and washed two times with PBS. They were next lysed using cold RIPA buffer (Thermo Scientific) supplemented with cOmplete protease inhibitor cocktail (Roche) for 30min, 4°C on rotation. Next, the samples were treated with 5µM dithiothreitol (DTT) and incubated at 56°C, 30min followed by alkylation using 10µM iodoacetamide (IAA) for 30min, RT in the dark. Proteins were precipitated in 90% EtOH over night at -20°C. The following day, the samples were centrifuged at 14000g, 4°C for 10min. The supernatant was discarded, and the pellets were dried using a SpeedVac. The samples were resuspended in 100µl of 0.1M ammonium carbonate buffer and the protein concentrations were determined at 280nm using a NanoDrop (DS-11 Series Spectrphotometer/Fluormeter, DeNovix) and trypsination (Sequencing grade modified trypsin, porcine, Promega) was performed (1:50 trypsin:protein ratio) overnight at 37°C. Next day, trypsin was inhibited with 0.4% trifluoroacetic acid (TFA) and the peptides were dried by SpeedVac and stored at -80°C until use.

The samples were resolved in 22µl 2% ACN and 0.1% TFA and peptide concentration were determined at 215nm using NanoDrop. The samples were diluted to 0.5µg/µl and 2µl was injected to Liquid chromatography mass spectrometry (LC-MS). The LC-MS detection was performed on Tribrid mass spectrometer Fusion equipped with a Nanospray Flex ion source and coupled with an EASY-nLC 1000 ultrahigh pressure liquid chromatography (UHPLC) pump (Thermo Fischer Scientific). Peptides were concentrated on an Acclaim PepMap 100 C18 precolumn (75μm x 2cm, Thermo Scientific, Waltham, MA) and then separated on an Acclaim PepMap RSLC column (75μm x 25cm, nanoViper, C18, 2μm, 100Å) at the temperature of 45°C and with a flow rate of 300nl/min. Solvent A (0.1% formic acid in water) and solvent B (0.1% formic acid in acetonitrile) were used to create a nonlinear gradient to elute the peptides. For the gradient, the percentage of solvent B was maintained at 3% for 3min, increased from 3% to 30% for 90min and then increased to 60% for 15min and then increased to 90% for 5min and then kept at 90% for another 7min to wash the column.

The Orbitrap Fusion was operated in the positive data-dependent acquisition (DDA) mode. The peptides were introduced into the LC-MS via stainless steel Nano-bore emitter (OD 150µm, ID 30µm) with the spray voltage of 2 kV and the capillary temperature was set to 275°C. Full MS survey scans from m/z 350-1350 with a resolution of 120,000 were performed in the Orbitrap detector. The automatic gain control (AGC) target was set to 4 × 10^5^ with an injection time of 50ms. The most intense ions (up to 20) with charge states 2-5 from the full scan MS were selected for fragmentation in the Orbitrap. The MS2 precursors were isolated with a quadrupole mass filter set to a width of 1.2m/z. Precursors were fragmented by high-energy collision dissociation (HCD) at a normalized collision energy (NCE) of 30%. The resolution was fixed at 30000 and for the MS/MS scans, the values for the AGC target and injection time were 5 × 10^4^ and 54ms, respectively. The duration of dynamic exclusion was set to 45s and the mass tolerance window was 10ppm.

The raw DDA data were analysed with Proteome Discoverer™ 2.5 Software (Thermo Fisher Scientific). Peptides were identified using SEQUEST HT against UniProtKB human database (SwissProt TaxID=9606_and_subtaxonomies). The search was performed with the following parameters applied: static modification: cysteine carbamidomethylation and dynamic modifications: N-terminal acetylation and methionine oxidation. Precursor tolerance was set to 15 ppm and fragment tolerance was set to 0.05ppm. Up to 2 missed cleavages were allowed and Percolator was used for peptide validation at a q-value of maximum 0.01. Extracted peptides were used to identify and quantify them by label-free relative quantification. The extracted chromatographic intensities were used to compare peptide abundance across samples. Protein abundances were normalized against total amount of peptides.

The data was subsequently analysed in Microsoft excel. The raw data is available via ProteomeXchange with identifier PXD033983. First, proteins identified with 0-2 unique peptides were excluded from further analysis. Next, data was filtered to include proteins found in ≥2 donors, and in ≥2 of the triplicates. The proteins fulfilling both criteria were used for statistical analysis, where the abundance values of the triplicates were averaged followed by paired t-test. In parallel, the values of the three donors were averaged, resulting in 2 groups, one for serum polarized monocytes and one for SF-polarized monocytes. These values were used to calculate the Log2 fold change. Proteins were considered upregulated if they had a log2 fold change of ≥1 and a p-value of <0.05, and downregulated if they had a log2 fold change of ≤ -1 and a p-value of <0.05.

Proteins that fulfilled the criteria (proteins found in ≥2 donors, and in ≥2 of the triplicates) for serum, but did not fulfil them for SF, were considered downregulated and combined with the proteins above. On the contrary, proteins that fulfilled the criteria for synovial fluid, but not serum, was considered upregulated and combined with the upregulated proteins. These proteins were then used for enrichment analysis (http://geneontology.org/) of altered biological processes using the PANTHER overrepresentation test (release 2022-02-02) and the GO Ontology database (DOI: 10.5281/zenodo.6399963, release 2022-03-22). Identified processes were considered if they had a false discovery rate adjusted p-value of <0.05 and ≥9 of the altered proteins included in the process. If several processes from the same gene ontology hierarchal tree were enriched, the most distal one with the lowest p-value was elected for analysis.

**Transwell Migration Assay**

Primary human fibroblast-like synoviocytes (FLS) from the knee (Cell Applications) were allowed to attach to the underside of 5µm pore-sized transwell inserts (Corning) in synoviocyte growth medium (Cell Applications) before the inserts were turned and placed in a 24-well plate (Corning) and cultured for 3-5 days. HMEC endothelial cells (ATCC) were subsequently added to the inside of the inserts and cultured in MCDB 131 medium (Gibco) supplemented with 10% fetal bovine serum, 10ng/ml hEGF, non-essential amino acids, sodium pyruvate and PenStrep for 72hrs before use.

The prepared inserts were placed in a new 24-well plate (Corning) with 400µl MCDB-131 medium supplemented with 20% synovial fluid (instead of 10% FBS). 0.2x10^6^ recently isolated monocytes from healthy donors (described above) were added to the inserts, and monocytes were allowed to migrate for 3hrs at 37°C, 5% CO_2_ followed by removal of the inserts. As a control, monocytes were instead added directly to wells containing the same medium and synovial fluids, but without inserts. The cells were incubated overnight at 37°C, 5% CO_2_. Next day, monocytes were detached, counted, and used for surface marker analysis (CD14, MerTK, CD86, HLA and CD16 (clone 3G8, APC-H7, BD) all diluted 1:200 and T-cell proliferation assays as described above.

**Co-Culture with Fibroblast-like Synoviocytes**

Primary healthy FLS were seeded in 96-well plates (Falcon) and grown for 72hrs in synoviocyte growth medium before use. Monocytes were isolated from healthy donors and 1x10^5^ cells, in 100µl RPMI-1640 medium containing 20% SF, were added to the FLS after two washes with RPMI-1640. Wells with monocytes and synovial fluid but without FLS served as controls. The cells were incubated overnight at 37°C, 5% CO_2_. Next day, monocytes were detached, counted, and used for surface marker analysis (CD14, MerTK, CD86, HLA and CD16 all diluted 1:200, and T-cell proliferation assays as described above).

**Statistics**

Flow cytometry data was analysed using the CytExpert software (v2.3) or Kaluza (v2.1, both Beckman Coulter). Data is presented as median with interquartile range if not otherwise stated. Paired data was analysed using the Wilcoxon matched pairs signed rank test and unpaired data with the Mann-Whitney U test. In cases of multiple comparisons, correction was performed using Bonferroni correction unless otherwise stated. The ratio of T-cell proliferation was calculated using one sample Wilcoxon signed-rank test and the hypothetical median of 1. Mass spectrometry data was analysed as described above. R programming language (3) was used to cluster the patients based on their joint markers into two groups by hierarchical clustering with “ward.D2” linkage function.  A random forest predictive model based on the blood markers estimated the importance of variables by evaluating mean decrease accuracy when dropping every variable from the model.  p<0.05 was considered statistically significant. All other data was analysed using GraphPad Prism 9 and Microsoft Excel.

3.	R Core Team. R: A Language and Environment for Statistical Computing. Vienna, Austria: R Foundation for Statistical Computing; 2022.

**Supplementary Figure legends**

**Supplementary Figure 1 The effect of tofacitinib and tocilizumab on monocyte viability is low** Monocytes were pre-treated with tocilizumab (100ng/ml) or tofacitinib (1µM) before being treated with a pool of synovial fluid (SF) from 6 oJIA patients and polarized overnight. Viability was assessed using Annexin V / PI staining and flow cytometry. Viable cells were defined as Annexin V / PI negative. Each condition was performed in triplicate, and the bars represent mean ±SD of 3 different monocyte donors. *oJIA- oligoarticular juvenile idiopathic arthritis, PI- Propidium iodide, tof- Tofacitinib, toc- Tocilizumab.*

**Supplementary Figure 2 Gating strategy used for monocytes in patients** (**A**) Representative gating strategy used for the analysis of surface markers of monocytes in synovial fluid (SF) and blood. (**B**) Representative gating strategy used for the analysis of STAT phosphorylation following activation in SF and blood. (**C**) Representative gating strategy for the analysis of intracellular cytokine production following LPS activation in SF and blood. *LPS- Lipopolysaccharide, IL- Interleukin, STAT- Signal transducer and activator of transcription, MFI- Median fluorescence intensity, TNF- Tumor necrosis factor.*

**Supplementary Figure 3 Generation of apoptotic neutrophils and gating strategy for efferocytosis** Neutrophils were isolated from healthy donors, stained with CTV, and incubated overnight in serum poor medium. (**A**) A representative experiment showing how apoptosis was confirmed for each experiment using Annexin V staining. (**B**) Representative gating strategy, showing serum and SF polarized monocytes which were gated as CD14^+^CD66b^-^ to exclude monocytes that bound, but did not internalize, apoptotic neutrophils. *CTV- Cell Trace Violet, SF- synovial fluid, Pol- Polarization.*

**Supplementary Figure 4 Strategy to identify down- and upregulated proteins identified by mass spectrometry in serum vs synovial fluid polarized healthy monocytes** Monocytes from n=3 healthy donors were polarized with a pool of serum or synovial fluid (SF) from patients with oJIA, and subsequently analysed by mass spectrometry. Proteins were next sorted and analysed, resulting in 66 downregulated proteins and 62 upregulated proteins, which were then used for gene ontology enrichment analysis of biological processes.

**Supplementary Figure 5 Migration induces some of the features observed in the patients’ monocyte phenotype** (**A**) Experimental setup of the migration assay, in which monocytes were allowed to migrate for 3hrs followed by removal of the inserts and polarization overnight (**B**) Non-migrated control (SF) and migrated (Mig) monocytes were detached and co-cultured with CTV-stained CD3 activated T-cells (1:10 monocytes to T-cells) for 72hrs, followed by analysis of proliferation (displayed as ratio of percent proliferation between migrated vs non-migrated monocytes) and (**C**) expression of activation markers in T-cells (n=22). (**D**) Shows changes in surface expression of CD86 and HLA in non-migrated vs migrated monocytes. (**E**) Displays ROS production after 1hr incubation following H_2_DCFDA staining (n=12) and (**F**) phagocytosis of opsonized FITC labelled beads for 30min (n=6). Wilcoxon matched pairs signed rank test. Lines at median. *HMEC- Human dermal microvascular endothelial cells, FLS- Fibroblast-like synoviocytes,* *ROS- Reactive oxygen species, MFI- Median fluorescence intensity, SF- Synovial fluid.*

**Supplementary Figure 6 Tofacitinib and tocilizumab do not inhibit the increased antigen presentation abilities by migrated or co-cultured monocytes** (**A**) Monocytes were pre-treated with tofacitinib (1µM) or tocilizumab (100ng/ml) before migration for 3hrs, followed by polarization overnight. Next day, they were analysed for expression of CD86 and HLA, or (**B**) detached and incubated with healthy CD3 activated T cells for 72hrs, which were analysed for proliferation and surface marker expression. (**C**) Instead of migration, monocytes were polarized and co-cultured with fibroblast-like synoviocytes overnight, and analysed for CD86, HLA or (**D**) T-cell activation. Wilcoxon matched pairs signed rank test, n=8. Lines at median with interquartile range. *MFI- Median fluorescence intensity, SF- Synovial fluid, FLS- Fibroblast-like synoviocytes, Tof- Tofacitinib, Toc- Tocilizumab, Mig- Migrated monocytes.*

**Supplementary Figure 7 Clinical features of the two patient groups** Clinical characteristics of patients categorized by hierarchical clustering based on IL-6/JAK/STAT into group one (low IL-6/JAK/STAT involvement, n=14) and group two (high IL-6/JAK/STAT involvement, n=19). (**A**) Shows percentage of the respective groups. (**B**) Displays the median with interquartile range. Statistics were performed with Mann-Whitney U test.
