## Supplementary figures and images for "Synovial Monocytes Drive the Pathogenesis in Oligoarticular Juvenile Idiopathic Arthritis via IL-6/JAK/STAT Signalling and Cell-Cell Interactions"

### Supplementary Figure 1

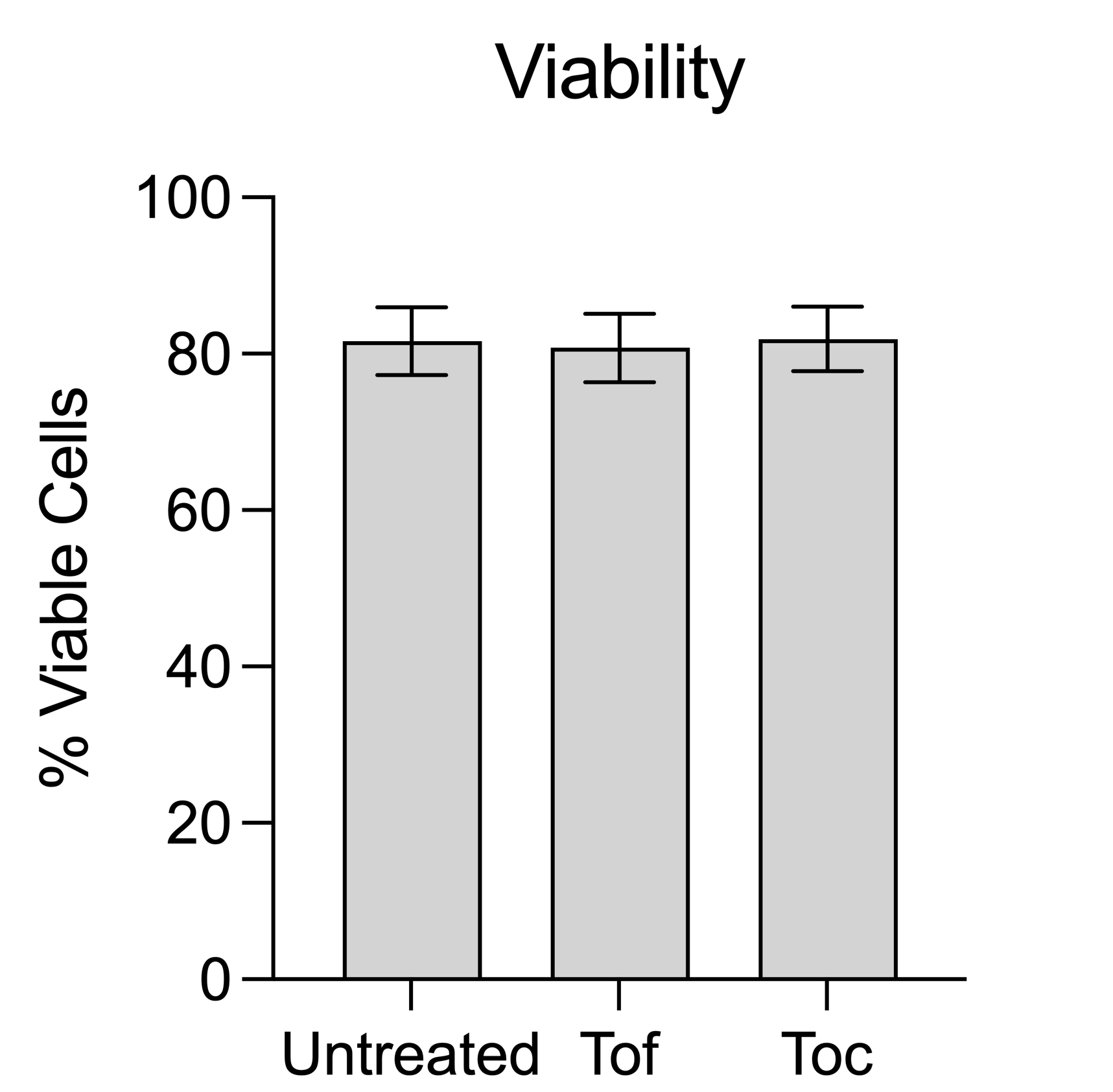

### Supplementary Figure 2

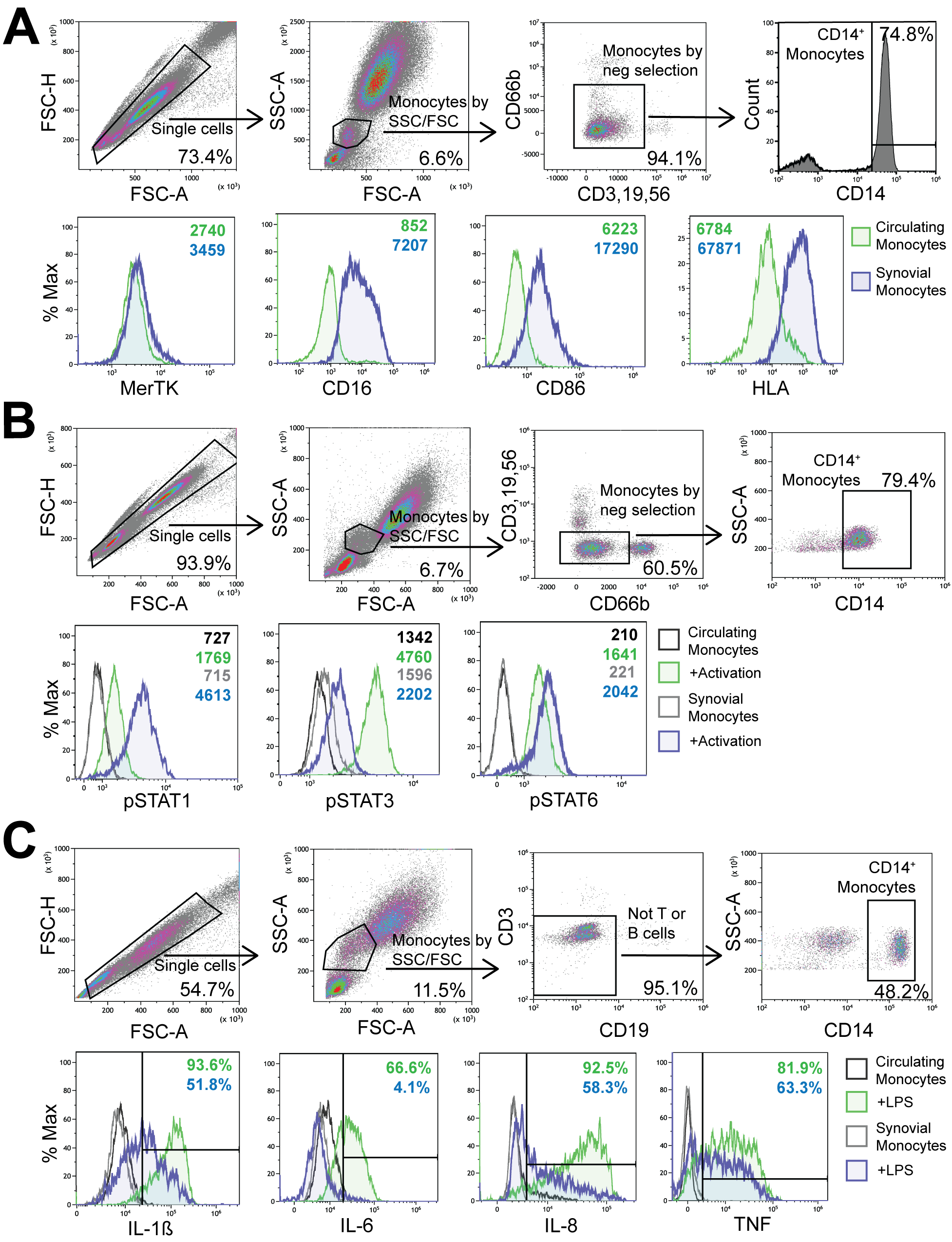

### Supplementary Figure 3

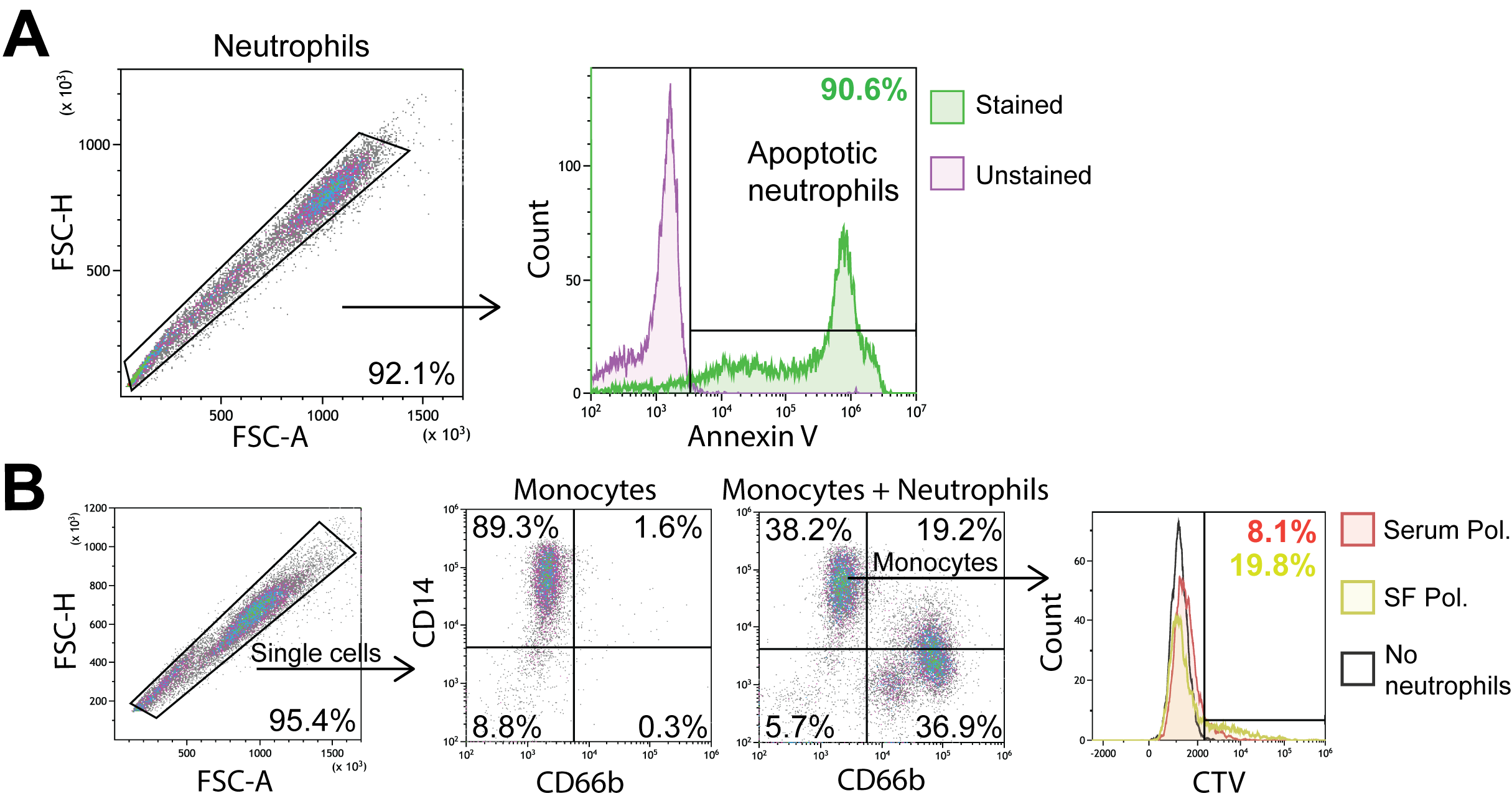

### Supplementary Figure 4

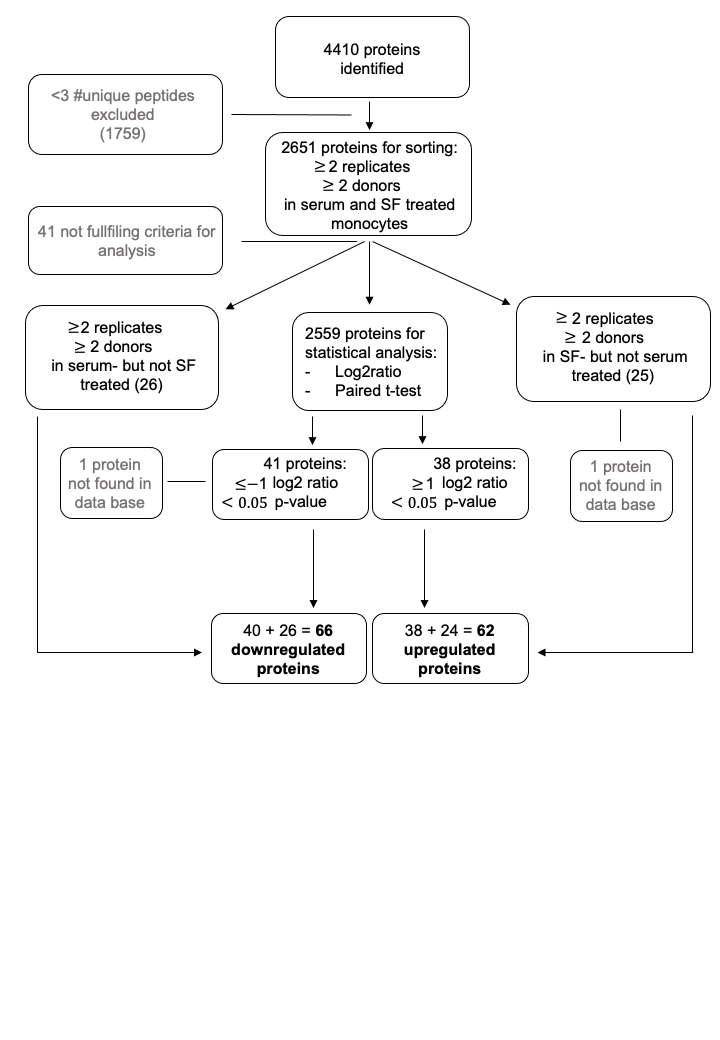

### Supplementary Figure 5

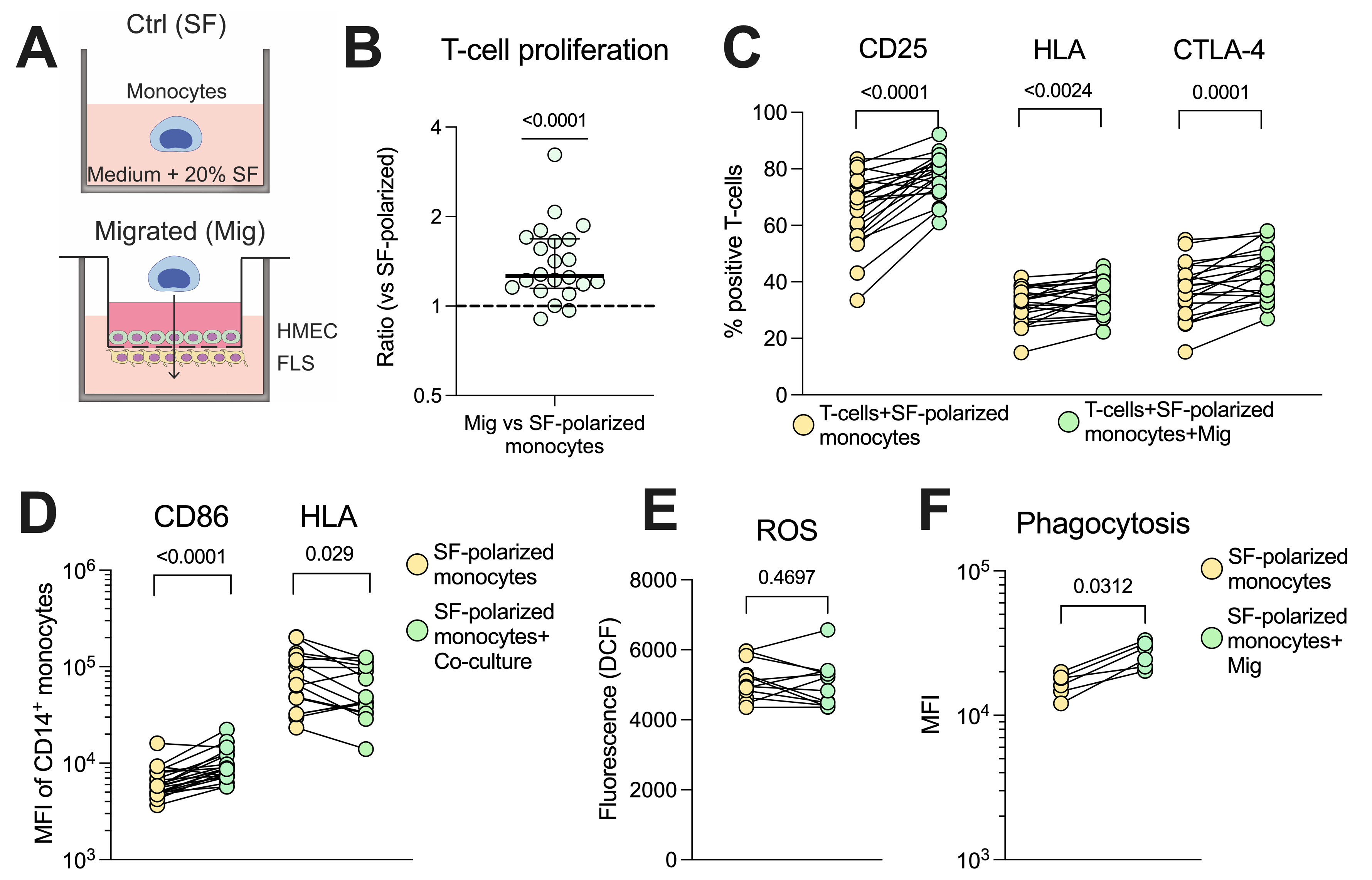

### Supplementary Figure 6

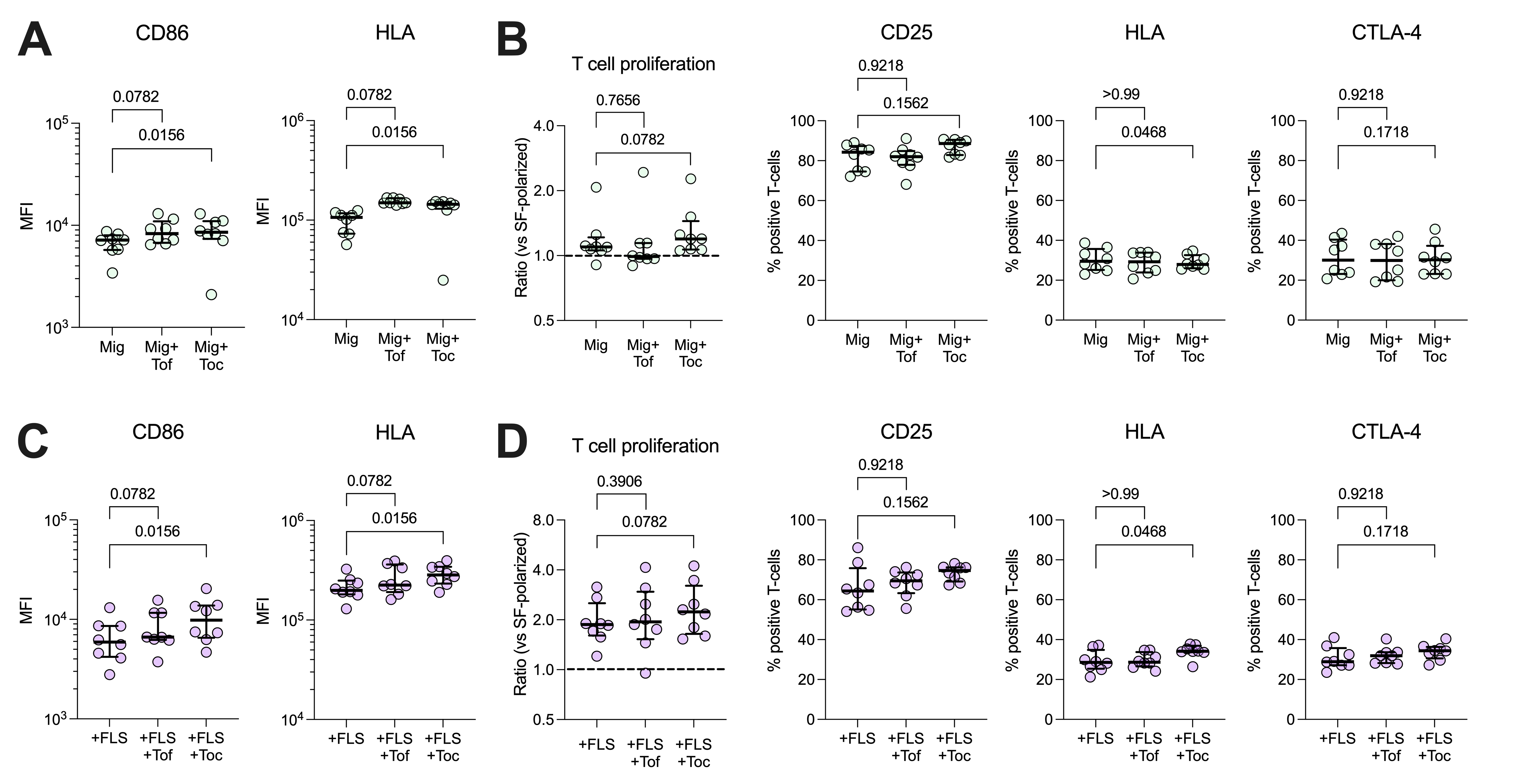

### Supplementary Figure 7

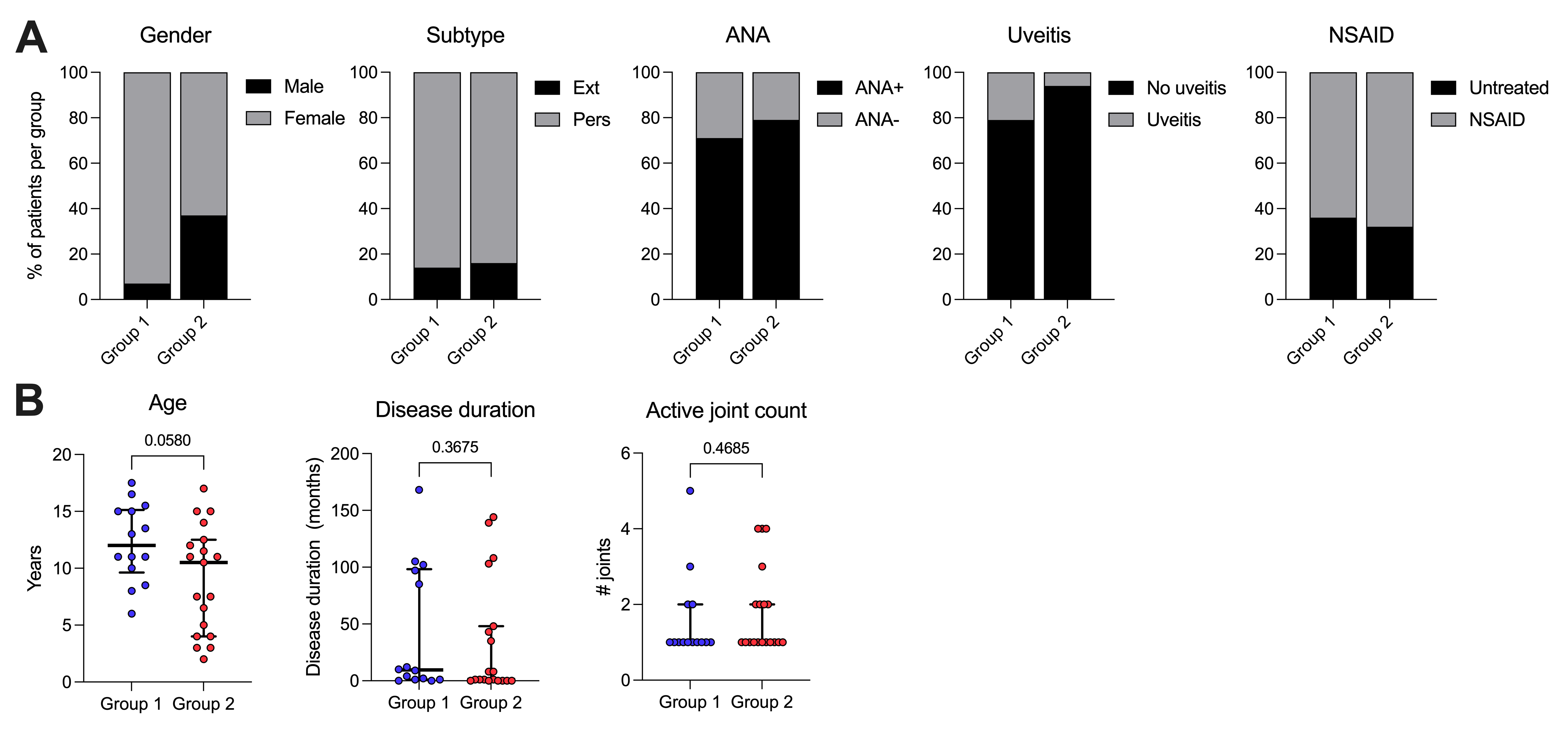
