## Supplementary Table 1 for "Synovial Monocytes Drive the Pathogenesis in Oligoarticular Juvenile Idiopathic Arthritis via IL-6/JAK/STAT Signalling and Cell-Cell Interactions"

**Supplementary table 1** Downregulated and upregulated proteins used for analysis of biological processes following liquid chromatography mass spectrometry.

| **Downregulated proteins** | | |  | | **Upregulated proteins** | | |
| --- | --- | --- | --- | --- | --- | --- | --- |
| **Entrez Gene ID** | **Gene Symbol** | | | **Entrez Gene ID** | | | **Gene Symbol** |
| 55937 | APOM | | | 80896 | | NPL | |
| 9859 | CEP170 | | | 1281 | | COL3A1 | |
| 22918 | CD93 | | | 2 | | A2M | |
| 7052 | TGM2 | | | 1667 | | DEFA1 | |
| 3726 | JUNB | | | 7045 | | TGFBI | |
| 6288 | SAA1 | | | 2312 | | FLG­ | |
| 7185 | TRAF1 | | | 6414 | | SELENOP | |
| 6291 | SAA4 | | | 55823 | | VPS11 | |
| 28820 | IGLV1-51 | | | 274 | | BIN1 | |
| 3732 | CD82 | | | 55191 | | NADSYN1 | |
| 23063 | WAPL | | | 2335 | | FN1 | |
| 10135 | NAMPT | | | 28448 | | IGHV3-15 | |
| 1051 | CEBPB | | | 9761 | | MLEC | |
| 2204 | FCAR | | | 23586 | | DDX58 | |
| 9765 | ZFYVE16 | | | 5538 | | PPT1 | |
| 6317 | SERPINB3 | | | 4644 | | MYO5A | |
| 23601 | | CLEC5A | | 2214 | | FCGR3A | |
| 3123 | | HLA-DRB1 | | 3113 | | HLA-DPA1 | |
| 3383 | | ICAM1 | | 3115 | | HLA-DPB1 | |
| 4791 | NFKB2 | | | 2219 | | FCN1 | |
| 84153 | RNASEH2C | | | 55340 | | GIMAP5 | |
| 1593 | CYP27A1 | | | 4026 | | LPP | |
| 79931 | TNIP3 | | | 9788 | | MTSS1 | |
| 319 | APOF | | | 3133 | | HLA-E | |
| 5055 | SERPINB2 | | | 11326 | | VSIG4 | |
| 84034 | EMILIN2 | | | 2752 | | GLUL | |
| 4170 | MCL1 | | | 2243 | | FGA | |
| 157769 | FAM91A1 | | | 2244 | | FGB | |
| 715 | C1R | | | 713 | | C1QB | |
| 10318 | TNIP1 | | | 972 | | CD74 | |
| 336 | APOA2 | | | 975 | | CD81 | |
| 5329 | PLAUR | | | 4176 | | MCM7 | |
| 6868 | ADAM17 | | | 3026 | | HABP2 | |
| 341 | APOC1 | | | 83666 | | PARP9 | |
| 79572 | ATP13A3 | | | 725 | | C4BPB | |
| 162394 | SLFN5 | | | 2517 | | FUCA1 | |
| 344 | APOC2 | | | 4312 | | MMP1 | |
| 5209 | PFKFB3 | | | 10457 | | GPNMB | |
| 10713 | USP39 | | | 4697 | | NDUFA4 | |
| 346 | APOC4 | | | 3929 | | LBP | |
| 23515 | MORC3 | | | 2266 | | FGG | |
| 126298 | IRGQ | | | 168537 | | GIMAP7 | |
| 348 | APOE | | | 5476 | | CTSA | |
| 7133 | TNFRSF1B | | | 1893 | | ECM1 | |
| 4318 | MMP9 | | | 1509 | | CTSD | |
| 3552 | IL1A | | | 64744 | | SMAP2 | |
| 3553 | IL1B | | | 51816 | | ADA2 | |
| 3045 | HBD | | | 8685 | | MARCO | |
| 51429 | SNX9 | | | 2158 | | F9 | |
| 3557 | IL1RN | | | 474344 | | GIMAP6 | |
| 490 | ATP2B1 | | | 5360 | | PLTP | |
| 28781 | IGLV5-45 | | | 2289 | | FKBP5 | |
| 8942 | KYNU | | | 27250 | | PDCD4 | |
| 5743 | PTGS2 | | | 8050 | | PDHX | |
| 23279 | NUP160 | | | 3698 | | ITIH2 | |
| 8943 | AP3D1 | | | 9332 | | CD163 | |
| 54386 | TERF2IP | | | 7412 | | VCAM1 | |
| 3827 | KNG1 | | | 3700 | | ITIH4 | |
| 2165 | F13B | | | 629 | | CFB | |
| 51703 | ACSL5 | | | 28410 | | IGHV3-72 | |
| 3576 | CXCL8 | | | 1277 | | COL1A1 | |
| 6520 | SLC3A2 | | | 23166 | | STAB1 | |
| 50809 | HP1BP3 | | |  | |  | |
| 55802 | DCP1A | | |  | |  | |
| 123 | PLIN2 | | |  | |  | |
| 8829 | NRP1 | | |  | |  | |
